# Risk-Averse Multi-Stage Stochastic Programming to Optimizing Vaccine Allocation and Treatment Logistics for Effective Epidemic Response

**DOI:** 10.1101/2021.05.28.21258003

**Authors:** Xuecheng Yin, İ. Esra Büyüktahtakın

## Abstract

Existing compartmental-logistics models in epidemics control are limited in terms of optimizing the allocation of vaccines and treatment resources under a risk-averse objective. In this paper, we present a data-driven, mean-risk, multi-stage, stochastic epidemics-vaccination-logistics model that evaluates various disease growth scenarios under the Conditional Value-at-Risk (CVaR) risk measure to optimize the distribution of treatment centers, resources, and vaccines, while minimizing the total expected number of infections, deaths, and close contacts of infected people under a limited budget. We integrate a new ring vaccination compartment into a Susceptible-Infected-Treated-Recovered-Funeral-Burial epidemics-logistics model. Our formulation involves uncertainty both in the vaccine supply and the disease transmission rate. Here, we also consider the risk of experiencing scenarios that lead to adverse outcomes in terms of the number of infected and dead people due to the epidemic. Combining the risk-neutral objective with a risk measure allows for a trade-off between the weighted expected impact of the outbreak and the expected risks associated with experiencing extremely disastrous scenarios. We incorporate human mobility into the model and develop a new method to estimate the migration rate between each region when data on migration rates is not available. We apply our multi-stage stochastic mixed-integer programming model to the case of controlling the 2018-2020 Ebola Virus Disease (EVD) in the Democratic Republic of the Congo (DRC) using real data. Our results show that increasing the risk-aversion by emphasizing potentially disastrous outbreak scenarios reduces the expected risk related to adverse scenarios at the price of the increased expected number of infections and deaths over all possible scenarios. We also find that isolating and treating infected individuals are the most efficient ways to slow the transmission of the disease, while vaccination is supplementary to primary interventions on reducing the number of infections. Furthermore, our analysis indicates that vaccine acceptance rates affect the optimal vaccine allocation only at the initial stages of the vaccine rollout under a tight vaccine supply.

## 1 Introduction

Epidemics and pandemics have devastated humanity throughout its existence. One recent example is the Coronavirus (COVID-19), which has spread all over the world since its first detection in China at the end of 2019, causing over 33 million cases and 1 million deaths as of October 2020 (John Hopkins University CSSE, 2020). The COVID-19 has also resulted in large economic losses, and the associated damage continues to escalate. For example, due to the pandemic, 400 million full-time jobs were lost across the world (CNBC, 2020), and consumer spending so far has decreased by more than one trillion dollars only in the U.S. (Routley, 2020). Another example is the 2014-16 Ebola Virus Disease (EVD), one of the deadliest viral infections, causing more than ten thousand deaths in West Africa. Other recent examples include the Severe Acute Respiratory Syndrome (SARS), which affected 26 countries since its discovery in South China in 2003, and the novel swine-origin influenza A (H1N1) virus that spread fast in the human population since its first appearance in 2009, causing tens of millions of cases and 12,469 deaths only in the U.S. (CDC, 2016, 2019; WHO, 2019c). Such viral diseases causing lower respiratory infections, such as pneumonia, have remained among the top causes of death globally, including stroke and cancer (Hasan et al., 2019; WHO, 2021b).

Effective and timely allocation of limited resources, such as medical treatment and vaccination, plays a crucial role in alleviating the ravaging impacts of infectious disease outbreaks on the human population. This problem has attracted much attention from academics and practitioners. The vast majority of the research literature involves simulations and differential equations (Ajelli et al., 2016; Craft et al., 2005; Kaplan et al., 2003; Siettos et al., 2015) to estimate the transmission of the disease and tackle the epidemic resource allocation problem. Other studies use network models and stochastic compartmental models to analyze various strategies on the control of an epidemic (Berman and Gavious, 2007; Funk et al., 2017; Lekone and Finkenstädt, 2006; Longini Jr et al., 2007; Porco et al., 2004; Riley and Ferguson, 2006; Tanner et al., 2008) as well as resource allocation analysis (Nguyen et al., 2017; Shaw and Schwartz, 2010; Tebbens and Thompson, 2009; Zaric et al., 2000).

Previous operations research models that study the epidemic diseases and resource allocation mainly focused on the logistics and operation management to control the disease in optimal ways (Büyüktahtakın et al., 2018; Ekici et al., 2013; Liu et al., 2019; Queiroz et al., 2020; Zaric and Brandeau, 2001). Only a few of those OR studies that integrate resource allocation with epidemics control consider the uncertain parameters for resource allocation to control the disease. Those mainly use stochastic and approximate dynamic programming (Coşgun and Büyüktahtakın, 2018; Long et al., 2018) and two-stage stochastic programming (Ren et al., 2013; Tanner et al., 2008; Yarmand et al., 2014). Because the growth of an infectious disease dynamically changes over time, Yin and Büyüktahtakın (2021) present a multi-stage stochastic programming model to capture the dynamics of an evolving disease for effective epidemic control under the uncertainty of disease transmission.

Multi-stage stochastic programs typically minimize an expectation criterion, which calculates the expected cost of all possible scenarios, each of which is mapped with a certain probability of occurrence. The expectation is the most widely-used objective criterion in stochastic programming (Ahmed, 2006). However, it does not capture the variability in possible scenarios that could arise, in particular, the situations with high impact and low probability. If some extreme scenarios occur, there could be a significant loss when only the expected value is considered in resource allocation decision-making. For example, in a disastrous epidemic outbreak situation, non-repetitive decisions made at the beginning of the horizon, such as the placement of treatment facilities, may result in capacity shortages and unmet demand under the realization of a severe disease spread scenario. At the beginning of an epidemic outbreak, disease characteristics, such as the infection or disease transmission rate, may be unknown, and the disease growth could be highly uncertain due to the lack of data. Thus a large number of infections and losses could happen in shorter time periods than expected, as in the case of COVID-19 (Lazzerini and Putoto, 2020; Li et al., 2020). The former epidemics control multi-stage stochastic programming model of Yin and Büyüktahtakın (2021) only considered an expectation criterion in the objective function. To alleviate the adverse impacts of experiencing a disastrous disease transmission scenario, we consider a risk measure in the objective function in addition to the expectation criterion.

Conditional value-at-risk (CVaR) is a coherent risk measure that can be used in an optimization model without losing convexity (Rockafellar and Uryasev, 2002). Therefore, many previous studies considered mean-risk models with CVaR in stochastic programming models (Ahmed, 2006; Miller and Ruszczyński, 2011; Rockafellar and Uryasev, 2002; Schultz and Tiedemann, 2006). CVaR-based mean-risk stochastic programming has been studied in various applications, such as supply chain management (Alem and Morabito, 2013), reverse logistic network design problem (Soleimani and Govindan, 2014), solid waste management system (Dai et al., 2014), water resources allocation (Zhang et al., 2016), and forestry invasive species control planning (Bushaj et al., 2020, 2021).

In this paper, we address the problem of building a mean-CVaR, multi-stage, stochastic mixed-integer programming epidemics-vaccination-logistics model. Our model evaluates various scenarios regarding the disease growth and the vaccine availability to optimize the distribution of treatment centers and vaccines while minimizing the total expected number of infections, funerals, and close contacts of infected people under a limited budget. Here, we consider the risk of experiencing scenarios that lead to adverse outcomes in terms of the number of infected and dead people due to the epidemic. Combining the risk-neutral objective with a risk measure allows for a trade-off between the weighted expected impact of the outbreak and the expected risks associated with experiencing extremely disastrous scenarios.

For a newly-discovered disease, the invention of a new vaccine is difficult and typically takes a long time. Even if there is an approved vaccine available, its production will be short compared to the high demand in the early period of the outbreak, and thus the availability of the vaccines will be limited. In this study, we address the optimal distribution of limited vaccine supply in addition to the allocation of treatment resources to control an epidemic outbreak under the uncertainty in the vaccine supply and the transmission rate.

To incorporate human mobility within multiple regions of a country, we present a new formulation to estimate migration rates among various locations. We apply our model to the case of controlling the 2018-2020 EVD in the Democratic Republic of the Congo (DRC). We provide insights into the optimal resource allocation among different regions of DRC in a multi-period planning horizon with and without risk. We also analyze how risk-aversion affects decision-making, such as the budget allocated to treatment and vaccination and the number of infections and deaths, compared to the risk-neutral problem.

## 2 Literature Review and Paper Contributions

This section presents a discussion of various approaches to epidemics control, logistics, and vaccination, focusing on the recent EVD outbreaks. We then give key contributions of this paper and managerial insights into the effective management of the EVD and similar disease outbreaks.

Statistical methods have been widely used to estimate the disease transmission parameters and forecast an infectious disease’s progression. For instance, Kelly et al. (2019) employed a non-parametrically estimated Hawkes point process model to generate multiple probabilistic projections of the 2018 − 2019 EVD outbreak size in DRC, while Dalziel et al. (2018) performed a retrospective analysis on community deaths during the 2014−2016 Ebola epidemic in Sierra Leone to estimate the number of unreported non-hospitalized cases.

Moreover, the majority of the literature has utilized compartmental models with simulations to study the strategies for controlling the EVD. For example, Jiang et al. (2017) constructed a modified epidemic model that takes hospital isolation, drug, and vaccines into account and later verified the result with a Monte Carlo Algorithm. Their work suggested that Ebola eradication requires systematic thinking, effective hospital isolation, and effective EVD drug use and vaccination. Mizumoto et al. (2019) presented a quantified effective reproduction number of the DRC’s ongoing EVD epidemic. Rachah (2018) proposed a deterministic compartmental model for assessing the impact of isolation to contain the EVD in Sierra Leone. According to their results, the isolation of latent detectable and infectious individuals is the most effective strategy in curtailing the virus.

Due to the high death rate and difficulties in Ebola treatment, new vaccinations have been developed and widely used to help control the disease. Various studies considered vaccination as a strategy and involved the uncertainty of the supply of vaccines. For instance, Kelly et al. (2019) used a stochastic branching process model to project the size and duration of the 2018 − 2019 Ebola outbreak in the Democratic Republic of Congo (DRC) under high (62%), low (44%), and zero (0%) estimates of vaccination coverage. Xie (2019) modified the Susceptible-Exposed-Infective-Hospitalized-Funeral-Removed model of Legrand et al. (2007) to examine disease transmission dynamics after vaccination for the 2014 Ebola outbreak in Liberia and found that the ring vaccination strategy would reduce the transmission rate. Chowell et al. (2019) employed an individual-level stochastic transmission model to evaluate ring and community vaccination strategies for the 2018 − 2019 DRC Ebola transmission. Their results also indicated that a ring vaccination strategy could speed up and enhance the probability of epidemic containment. Brettin et al. (2018) constructed a game-theoretic model of the EVD incorporating individual decisions on vaccination to study the effect of a promising Ebola vaccine (rVSV-ZEBOV). Their result showed that Ebola could be eradicated if voluntary vaccination programs are coupled with focused public education efforts.

Area et al. (2017) used simulations to compare compartmental models with and without vaccination. They concluded that vaccination of all susceptible individuals at the beginning of the outbreak would give the best result for controlling Ebola, and satisfactory results could be attained if the number of available vaccines meets the population’s needs. Wells et al. (2019) presented a method that can be applied to identify areas at risk during the 2019 EVD case in the DRC. They used a spatial model that incorporates human mobility, poverty, and population density and assessed the effectiveness of the vaccination. Their results demonstrated that even modest delays in initiating vaccination would have noticeably degraded the impact of the program.

Büyüktahtakın et al. (2018) developed an epidemics-logistics model to address the resource allocation problem for controlling an infectious disease outbreak and demonstrated the model in the case of 2014-15 EVD in West Africa. Their optimization model simultaneously considered the logistics problem and the disease growth and represented the transitions from the infections to the treatment compartment as a variable since this transition depends on the treatment center’s capacity. Yin and Büyüktahtakın (2021) extended the deterministic epidemics-logistics model of Büyüktahtakın et al. (2018) to a multi-stage stochastic mixed-integer programming model. They introduced the value of the stochastic solution (VSS) to study the advantages of the stochastic model compared to the deterministic model. They defined equity constraints to address the fair resource allocation problem on the logistics of epidemic control. They implemented the model on the 2014 EVD in West Africa, and their findings suggested that the multi-stage stochastic model is superior to the deterministic model, and forcing equity resource allocation too much would cause a significant loss on controlling the disease.

### 2.1 Key Contributions of the Paper

Former studies on the logistics of epidemics have omitted the risk of experiencing extreme scenarios when formulating a stochastic optimization model. A risk-neutral stochastic programming approach, which does not consider variability in possible scenario outcomes, may perform poorly when there are outliers in the distribution of the scenarios. Also, existing mathematical programming studies on epidemic control have not incorporated vaccine allocation into a compartmental-logistics model. Furthermore, due to the lack of data, the rates of migration among multiple regions of DRC are not known. The population is quite mobile within regions and countries in Africa (Flahaux and De Haas, 2016). Thus, movement rates are difficult to estimate.

In this paper, we address those aforementioned limitations existing in both the epidemiological modeling and healthcare operations research literature. Below, we present the modeling and applied contributions with key recommendations to decision makers.

#### Modeling Contributions

First, to our knowledge, we present the first risk-averse multistage stochastic programming model presented in the research field of infectious disease control. Different than the former literature, we formulate the uncertainty in the transmission rate from the close contacts of infected people to the infections compartment and the uncertainty in total vaccines available as two dependent random variables in a multi-stage stochastic scenario tree. We then incorporate a nested CVAR risk measure into the objective function of the formulation while defining risk-related constraints to alleviate the risk of experiencing scenarios that lead to adverse outcomes in terms of the number of infected and dead people due to the epidemic. We also provide insights on how the expected impact and expected risk in terms of deaths and infections change as the decision maker shifts from being risk-neutral to risk-averse at varying risk levels.

Second, we address the optimal allocation of vaccines to multiple regions within a country in addition to the allocation of Ebola Treatment Centers (ETCs) and treatment resources to control an epidemic outbreak in a multi-stage stochastic mean-risk model. Specifically, we have extended the Susceptible-Infected-Treated-Recovered-Funeral-Burial epidemics-logistics model of Büyüktahtakın et al. (2018) into an epidemic-vaccination-logistics model by incorporating a new ring vaccination compartment under uncertainty and risk.

Third, we develop a new formulation to estimate the migration rates between regions of a country and integrate the impacts of human mobility into our epidemic-vaccination-logistics model. Thus, our mathematical model captures the influence of human movement on the transmission of the disease.

Fourth, our risk-averse epidemic-vaccination-logistics model is general and thus could be adopted to study other epidemic diseases, such as influenza and H1N1, as well as pandemics, such as the COVID-19.

#### Applied Contributions and Key Recommendations to Decision Makers

We implement our multi-stage stochastic mean-risk model to study the case of the 2018-2020 EVD in the DRC.We collect and synthesize epidemiological, population, and economic data of Ebola infections in the provinces of DRC and organize them into regional data, using WHO Ebola situation reports (WHO, 2021a, 2020). We perform computational experiments to analyze the impact of treatment budget, risk parameters, uncertain vaccine availability, and vaccine acceptance and effectiveness rates on the allocation of resources, such as ETC and vaccines, during an epidemic. We drive several insights into the optimal resource allocation under various risk levels that the decision maker is willing to take for controlling an infectious disease. As such, our mathematical model could be used as a decision support tool to aid policymakers in determining the optimal risk-averse treatment and vaccine-allocation policies.

Based on our results, we provide the following recommendations to inform the resource allocation decision making under an epidemic situation:

i. Regions with a high initial infection level (“the number of infected people in a region” / “the total number of infected people” - “population in a region” / “total population over all regions”) get the majority of the resources. While the ETCs and treatment budget are mainly allocated to highly infected locations, the model allocates a budget for vaccination to most locations to prevent the disease’s spread. Our findings also suggest using the budget for vaccination in regions where the disease has just started, while in regions with high initial infections, the model gives priority to build new ETCs and treat infected people over vaccination.
ii. The potential risk associated with regions with low or zero initial infection levels should also be taken into account when making resource allocation decisions. For instance, a non-infected region nearby a highly-infected location may also be severely affected by the disease due to human mobility among multiple regions. Thus, as the risk-averseness level increases, the budget allocated to areas with the highest initial infection level is decreased by moving the ETC and treatment budget to neighboring locations under the risk of getting infections.
iii. A risk-averse decision-maker should expect a possible increase in the number of infections and deaths while trying to mitigate disastrous outbreak scenarios. Being risk-averse also increases the expected cost of treatment and vaccination.
iv. For the considered case of the EVD in DRC, isolating and treating infected individuals are the most efficient ways to slow the disease’s transmission. When the supplied vaccines are available but limited, the vaccination is supplementary to the primary interventions on reducing the number of infections.
v. While vaccination is supplementary, its delay could cause an exponential increase in the number of infections and deaths, even under the main intervention measures, such as treatment and isolation. In particular, vaccination at earlier stages of an epidemic would also help control the disease faster than immunization at later stages. Thus, if available, vaccination should be applied as early as possible for effective epidemic response.
vi. The number of vaccines supplied to Upper North Kivu and Middle North Kivu, the two most-impacted regions of DRC, has a complementary relationship. When the vaccine acceptance rate is fixed, the number of vaccines provided to these two regions only sightly fluctuates under different vaccine effectiveness rates. Also, the more effective the vaccine is, the fewer vaccines are needed in highly-impacted areas so that some remaining vaccines could be used in regions with lower infection.
vii. When the vaccine effectiveness rate is fixed, vaccine acceptance rates affect vaccine allocation at the initial stages of the vaccine rollout. Under a very low vaccine acceptance rate, available vaccines are moved from highly impacted locations to less affected areas. However, vaccine acceptance rates do not impact the total number of vaccines distributed throughout the planning horizon under a limited vaccine supply.

## 3 Multi-Stage Risk and Time Consistency

Let *F*_*Z*_(·) be the cumulative distribution function of a random variable *Z*. The *α*-quantile of the distribution, inf_*η*_{*η* ∈ℝ : *F*_*Z*_(*η*) ≥ *α*}, is defined as the value-at-risk (VaR) at the confidence level *α* ∈ [0, 1) and denoted by VaR_*α*_(*Z*).

The mean excess loss or tail VaR, at level *α*, is called conditional value-at-risk (CVaR), defined as CVaR_*α*_(*Z*) = 𝔼 (*Z* | *Z* ≥ VaR_*α*_(*Z*)). Specifically, CVaR is the conditional expected value that exceeds the VaR at the confidence level *α*. For a minimization problem, VaR_*α*_ is the *α*-quantile of the cost distribution, and it provides an upper bound on the cost that is exceeded only with a small probability of 1 − *α*. On the other hand, CVaR_*α*_ measures an expectation of the cost that is more than VaR_*α*_, the *α*-quantile of the distribution of costs. The conditional value-at-risk can be calculated as an optimization problem as follows (Rockafellar and Uryasev, 2002):

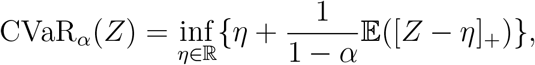

where (*a*)_+_ := *max*(*a*, 0) for any *a* ∈ℝ.

In this paper, we study a mean-risk minimization problem, as first introduced in Markowitz (1991):

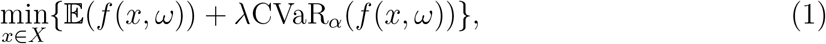

where 𝔼 (*f* (*x, ω*)) represents the expected cost function of the scenarios *ω* ∈ Ω, CVaR_*α*_ represents the conditional value-at-risk at *α* ∈ [0, 1), and *λ* ∈ [0, 1] is a non-negative coefficient of the risk part. The risk preference parameter *λ* is the weight of the risk term in the objective function (1), and can be adjusted for a trade-off between optimizing an expectation value [𝔼 (*f* (*x, ω*))] and the level of risk taken [CVaR_*α*_(*f* (*x, ω*))]. The larger the *λ*, the more risk-averse the decision maker is. The *α* parameter, on the other hand, gives the confidence level on the perceived risky scenarios that exceed the maximum acceptable loss, VaR_*α*_. As *α* increases, the probability of exceeding the VaR_*α*_ reduces, and thus the decision maker becomes more risk averse. The parameter *α* ∈ [0, 1) is typically set to a high value, e.g., 0.95. The parameters *λ* and *α* are set by the user to adjust the level of risk averseness and do not have a direct relationship.

### Time Consistency

When modeling a risk-averse multi-stage stochastic program, time consistency is considered as a critical issue. Time consistency implies that if you solve a multi-stage stochastic programming model today and find solutions for each node of a tree, you should get the same solution if you resolve the problem tomorrow when you are given the information that is observed and decided today. For a multi-stage stochastic model, risk measures can be applied at every stage additively or to the complete scenario path or in a nested form similar to dynamic programming. The nested risk measures are shown to satisfy the time consistency of multi-stage stochastic programs in the study of (Homem-de Mello and Pagnoncelli, 2016).

We consider a nested risk measure, expected conditional value-at-risk (𝔼-CVaR), as defined in Homem-de Mello and Pagnoncelli (2016). The 𝔼-CVaR can be linearized and formulated as a linear stochastic programming model. In the next section, we will utilize the 𝔼-CVaR as a risk measure to formulate our mean-risk multi-stage stochastic epidemics-vaccination-logistics model.

## 4 Problem Formulation

This section presents the compartmental model description and the mean-risk formulation of the multi-stage stochastic epidemics-vaccination-logistics model. In Appendix A, we describe the model notation that will be used throughout the rest of this paper.

### 4.1 Compartmental Disease Model Description

Figure 1 shows the transmission dynamics of the EVD in a region *r* located in DRC for each period *j*. The disease spreads among susceptible individuals (S) as well as close contacts of infected (H), by either person-to-person contact at a periodic rate of 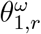 under scenario *ω* or through touching Ebola-related dead bodies that are not yet buried during traditional funerals at a periodic rate of *θ*_2,*r*_ in region *r*. Thus, once close contacts (H) or susceptible individuals (S) become infected, they move to the infected (I) compartment. However, individuals in the general community (S) are infected with a lower rate of *σ*_*r*_ compared to close contacts (H).

**Figure 1:**
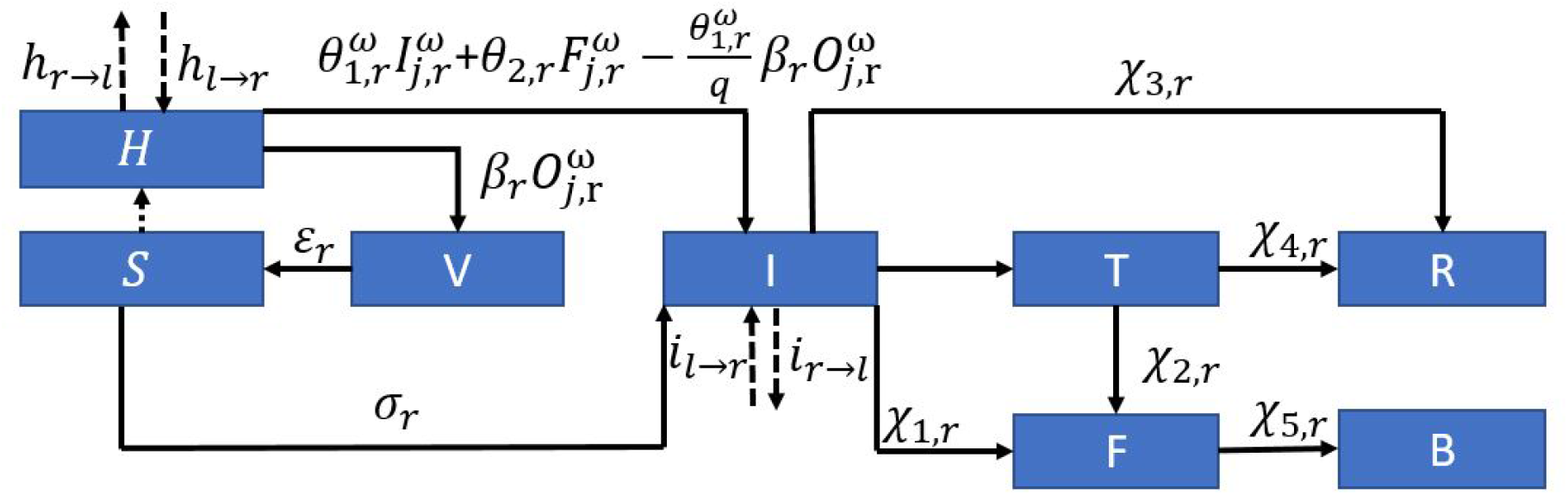
One-Step Disease Compartmental Model.

Here, we focus on modeling the ring vaccination, where only close contacts with infected people can be vaccinated. The quantity of vaccines allocated to region *r* under scenario *ω* at the end of period *j* under scenario *ω* is defined by the variable 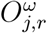. Given the effectiveness rate of vaccination, *β*_*r*_, there will be 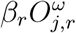 people who are moving from the (H) to (V) compartment in each period *j*, where (V) represents the successfully vaccinated individuals, who become fully immunized by vaccination. Next, due to the time effect, successfully vaccinated individuals (V) become no longer immune to the disease over time and move to the general community (S) with a rate of *ε*_*r*_ in region *r*. When the number of infected people increases, the close contacts of infected people will also increase. Thus, the dotted arrow from the general community (S) to close contacts (H) represent the movement of people from (S) to (H) as (I) increases. Furthermore, since people are getting vaccinated, the number of close contacts in (H) being infected and moving into the (I) compartment should be decreased by 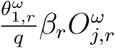, where *q* is the average number of close contacts for each infected individual. The term 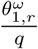 shows the proportion of close contacts that would be infected for each infected individual, and because 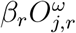 represents the number of successfully vaccinated close contacts who become fully immunized by vaccination, 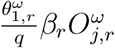 gives the number of close contacts saved by the vaccination.

Without treatment, some of the infected individuals in the compartment (I) will die and move to the funeral (F) compartment with the rate of *χ*_1,*r*_, while some of the infected individuals will recover with a rate of *χ*_3,*r*_, moving into the recovered compartment (R). However, the number of individuals hospitalized for treatment (T) is based on the treatment capacity variable 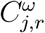, which gives the available number of beds in the ETCs in region *r* under scenario *ω* in period *j*. Thus, there is no constant transition rate from I to T. Meanwhile, individuals who did not receive treatment will remain in the community and spread the disease. In treated compartment (T), some individuals will recover with a periodic rate of *χ*_4,*r*_, and a fraction of them will die with a periodic rate of *χ*_2,*r*_. The deceased individuals in the funeral compartment are safely buried at a rate of *χ*_5,*r*_, moving into the buried compartment (B).

To describe the migration of infected individuals and close contacts within a given country, we define *i*_*l*→*r*_ and *i*_*r*→*l*_ as the rates of migration of infected individuals into and out of region *r*, as shown in dotted arrows going in and out of compartment (I). Similarly, we define *h*_*l*→*r*_ and *h*_*r*→*l*_ as the rates of migration of close contacts of infected people (H) into and out of region *r*.

### 4.2 Uncertainty and Model Assumptions

#### Modeling Uncertainty

In this paper, we used a discrete set of scenarios *ω* ∈ Ω to model the uncertainties related to the disease–the uncertainty in the transmission rate from the close contacts of the infected individuals to infections and the uncertainty in total vaccines supplied (available) at each stage *j*. Each scenario has a probability of *p*^*ω*^, where 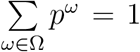. We assume that the uncertainty of the transmission rate is highly dependent on the availability of vaccine supply. If the vaccine supply is high, we observe a low transmission rate from close contacts to infections, and if the vaccine supply is low, we will have a high transmission rate instead. For our multi-stage stochastic model, we have two branches in each node of the scenario tree, representing the two possible realizations in each stage *j*: low and high transmission rates 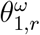 corresponding to the high and low levels of vaccines supplied at time *j* under scenario *ω*, 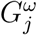. The case study values of the uncertain transmission and the vaccine availability parameters under two realizations at each branch of the scenario tree are presented in Table 10 in Appendix B.4.

#### Assumptions on Model, Data, and Parameters

The transmission of the EVD is affected by many factors, including damaged public health infrastructures, cultural beliefs, behavioral practices, and violent events frequently happening in DRC (Wannier et al., 2019; WHO, 2015). Due to these issues, data to calibrate some of the model parameters is either lacking or inaccurate. Data parameters, such as the transmission rate and the probability of scenarios, are quite difficult to estimate. Therefore, we make assumptions about some of the parameters used in the model formulation.

First, in our current model, each infected individual is assumed to have 100 close contacts, including direct close contacts and their close contacts (i.e., contacts of contacts) (CDC, 2015; Doshi et al., 2020). Those close contacts (e.g., household and health care workers) are considered as the high-risk group to be infected. In contrast, the susceptible people in the general community are considered in the lower risk group compared to infected people’s close contacts. Due to the nature of the ring vaccination, we assume that only the close contacts of infected people will get the vaccination.

Second, in our model, each node of the scenario tree has two realizations of the transmission rate and vaccine availability, as low and high. Former research has shown that violent events happening in DRC contribute to the increased transmission of EVD (Wannier et al., 2019). During our planning horizon of 15 weeks, from June 25, 2019, to October 8, 2019, seven violent events were reported (Dickey, 2018). Considering the likelihood of a violent event happening in each period, we assign a probability of 0.5 to each potential outcome (low and high) of disease transmission in our scenario tree in each time period. Disease transmission rates under no violent events are reported to vary between 0.81 and 1.08 (Wannier et al., 2019). Thus, we use a low transmission rate of 0.948 for North Kivu and 0.84 for Ituri based on the estimations reported in Camacho et al. (2014) and Wannier et al. (2019). The mean value of the transmission rate in DRC is 1.11 (Wannier et al., 2019), and the highest transmission rate in history is 1.83 (Chowell et al., 2004). In between those two values of transmission rates in DRC, we consider a high transmission rate of 1.422 for North Kivu and adjust it proportionally to a high transmission rate of 1.26 for Ituri based on the ratios of low transmission rates in both locations. Similar to their impacts on transmission rates, we assume that violent events lead to a low vaccine supply upper bound due to hindered humanitarian operations and lowered access to the infected population.

Third, the transmission rate defined in our model represents how many new infections can be generated from the existed infected individuals through a specific period of time. Under a small population, the transmission rate may be influenced by the total number of susceptible individuals since the total number of infected individuals may approach the maximum population size. However, here, we focus on meta-population modeling over multiple regions of a country. Thus the size of the susceptible population will not decline enough to make a significant difference in the infection rate for Ebola on a large spatial scale under several interventions already in place. Formulating the dependency of the susceptible population in the general community and the number of newly infected individuals also results in non-linearity in the optimization model. Thus, to avoid this non-linearity, we assume that the number of newly infected individuals in our model is independent of the size of the susceptible population in the general community.

Finally, current Ebola vaccines are shown to provide immunization for at least two years with high and stable levels of antibodies to the Ebola Zaire Virus in the blood of volunteers who are vaccinated (Branswell, 2018; WHO, 2021a). Thus, we assume that recovered individuals will not get infected again within four months of vaccination or recovering from the disease, which is nearly the planning horizon we consider in our study.

### 4.3 Model Formulation

Using the notation defined in Appendix A, the mean-risk multi-stage stochastic epidemic-vaccination-logistics model can be formulated as follows:

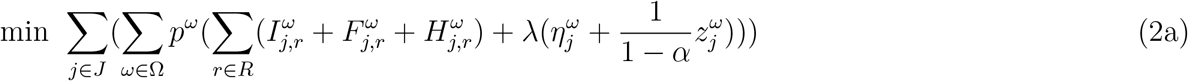

s.t. *Initial Condition Constraints* :

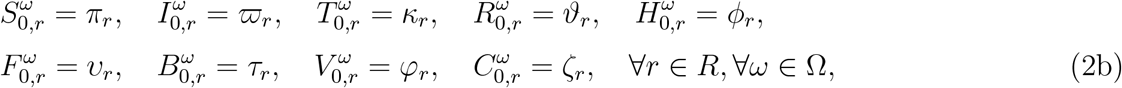

*Population Dynamics Constraints* :

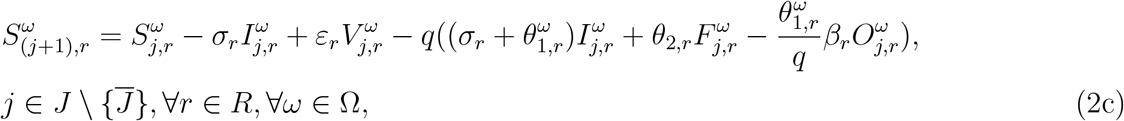

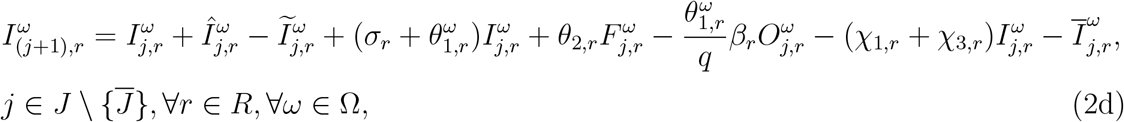

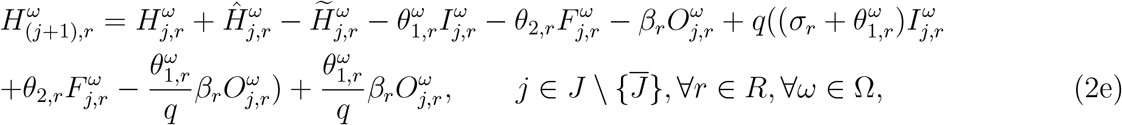

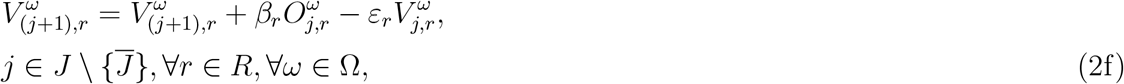

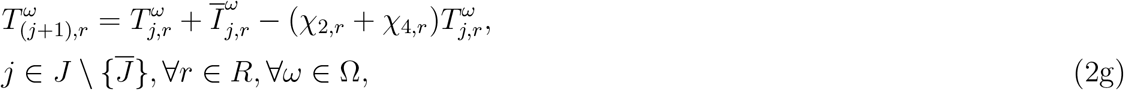

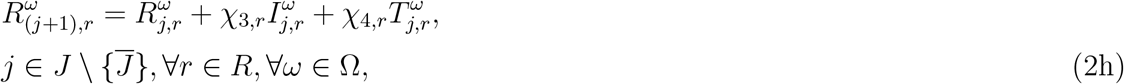

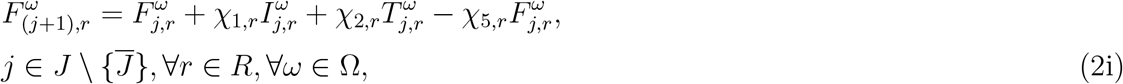

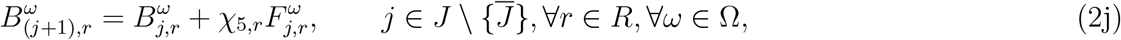

*Migration Constraints* :

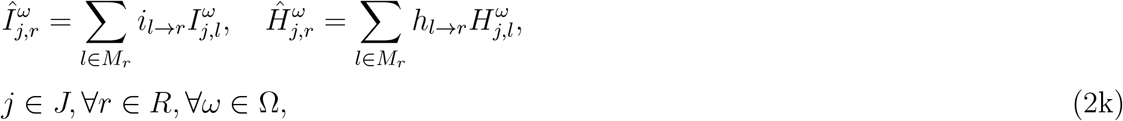

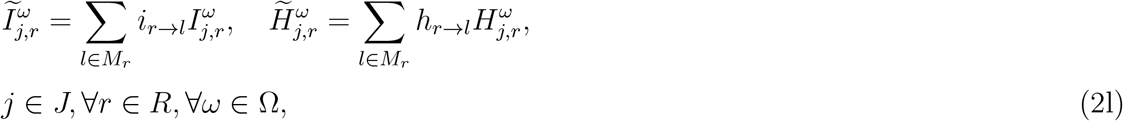

*Logistics and Operation Management Constraints* :

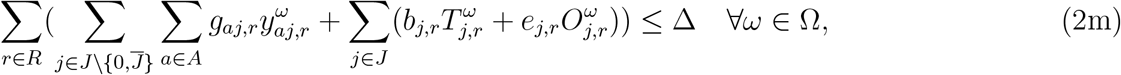

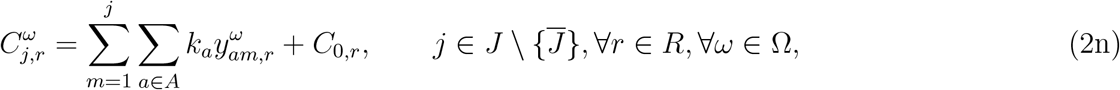

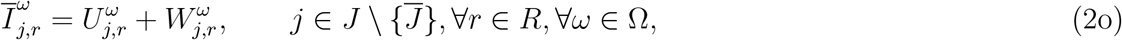

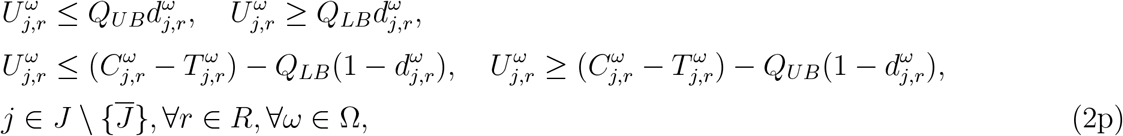

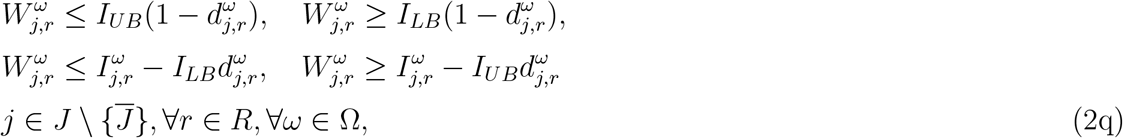

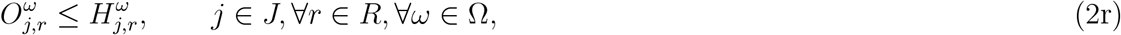

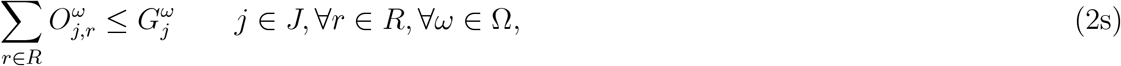

*Risk Measure Constraints* :

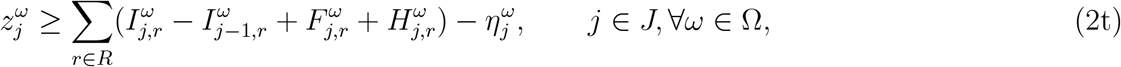

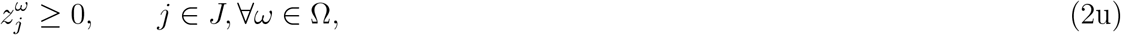

*Non* − *anticipativity Constraints* :

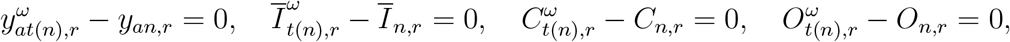

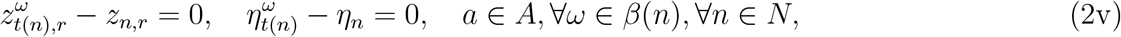

*Non* − *negativity Constraints* :

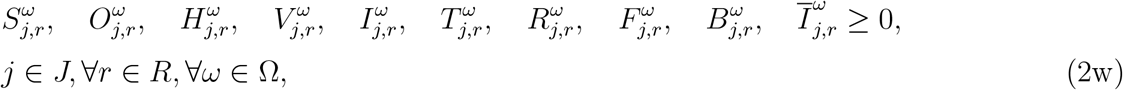

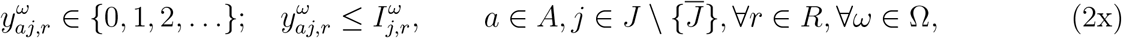

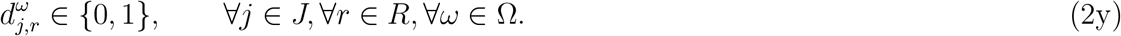

The objective function (2a) minimizes the total expected number of infected individuals, funerals, plus the close contacts of infected individuals, plus the conditional value-at-risk over all scenarios, in all regions throughout the planning horizon.

#### Initial Condition Constraints

Constraints (2b) give the initial number of the general community, infected, treated, recovered, close contacts, funerals, buried, and vaccinated compartments, and the total ETC capacity, respectively, in each region *r* at the beginning of the planning horizon.

#### Population Dynamics Constraints

Equations (2c)–(2j) represent the dynamics of the population in each disease compartment, which are shown in Figure 1. Constraint (2c) shows that the number of susceptible individuals in the community in region *r* at the end of period *j* + 1 under scenario *ω* is equal to the number of susceptible individuals from the previous time period minus the number of newly infected individuals, plus the number of successfully vaccinated individuals who were no longer immune to the disease at the end of period *j* under scenario *ω*. Furthermore, the term 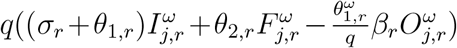 represents the number of susceptible individuals in the community (S) that transfer to close contacts (H) due to the newly infected individuals (I), where *q* is the average number of close contacts of each newly infected individual.

Constraint (2d) implies that the number of infected individuals at the end of period *j* + 1 in region *r* under scenario *ω* is equal to the number of infected individuals from the previous time period plus the net migrated infected individuals, plus newly infected individuals from the close contacts, general community, and funerals, minus individuals who were saved from infection by vaccination 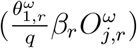 minus recovered, died, and treated individuals at the end of period *j* under scenario *ω*.

Constraints (2e) define the number of close contacts at the end of time period *j* + 1, which equals the number of close contacts from the previous time period plus the net migrated close contacts, minus the number of newly infected individuals from close contacts, minus the number of successfully vaccinated individuals during period *j*, and plus the new close contacts and the close contacts saved by the vaccination. Constraint (2f) ensures that the number of successfully vaccinated individuals at the end of period *j* +1 equals the number of successfully vaccinated individuals from the previous time period plus the number of new successfully vaccinated individuals and minus the number of individuals who are not immune to the virus anymore. Constraint (2g) describes the total number of treated individuals in region *r* at the end of time period *j* + 1 under scenario *ω*, which is equal to the number of treated individuals at the end of period *j* plus infected individuals who were admitted to the hospital for treatment based on the availability of beds minus treated individuals who died or recovered. Constraint (2h) implies that the cumulative number of individuals who recover in region *r* at the end of the period *j* + 1 under scenario *ω* is equal to the number of individuals who recover from the previous period plus newly recovered individuals. Constraint (2i) ensures the number of unburied funerals in region *r* at the end of time period *j* + 1 under scenario *ω* is equal to the sum of infected and treated individuals who died, minus the buried dead people. Constraint (2j) gives the total number of buried dead bodies at the end of the period *j* under scenario *ω*.

#### Migration Constraints

Constraints (2k) and (2l) formulate the number of net immigrated individuals in infected and close contact compartments, similar to the spatio-temporal reaction-diffusion (RD) models (Kıbış and Büyüktahtakın, 2019; Kıbış et al., 2020). Specifically, constraints (2k) show the number of infected individuals and close contacts migrating into region *r* from region *l* ∈ *M*_*r*_ under scenario *ω*. Constraints (2l) represent the number of infected individuals and close contacts emigrating from region *r* into neighboring region *l* ∈ *M*_*r*_ under scenario *ω*.

#### Logistics and Operation Management Constraints

Constraints (2m)–(2s) show the restrictions regarding logistics and operations management. Specifically, the inequality (2m) represents the budget constraint on the sum of the fixed costs of opening ETCs and the variable cost of treating infected individuals, and the cost of allocating vaccines over all regions *r* in all periods *j* under scenario *ω*. Constraint (2n) denotes the total capacity in region *r* at the end of period *j* under scenario *ω*. Constraint (2o)–(2q) are linear constraints that ensure the number of available beds in ETCs limit the number of hospitalized individuals in region *r*. Particularly, linear equations (2o)–(2q) are equivalent to the non-linear constraints implying that the number of hospitalized individuals (*Ī*) is equal to the minimum number of infected individuals and the capacity available at established ETCs after considering currently hospitalized individuals in ETCs (see Yin and Büyüktahtakın (2021) for the details of the linearization). Constraint (2r) represents that the number of vaccines supplied to region *r* at period *j* under scenario *ω* is limited by the number of close contacts in region *r* at period *j* under scenario *ω*. Constraint (2s) ensures that the total number of vaccines allocated over all regions can not exceed the available supply at each time period.

#### Risk Measure Constraints

Constraints (2t) and (2u) represent the risk measure limitations. Constraint (2t) calculates the difference between the objective function value and the value-at-risk for each stage under each scenario. Constraint (2u) ensures that the loss value exceeding the value-at-risk is included in the CVaR calculation, and thus 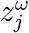 should be greater than or equal to 0.

#### Non-anticipativity Constraints

Constraints (2v) are non-anticipativity restrictions stating that if two scenarios share the same path up to stage *j*, the corresponding decisions will be the same.

#### Non-negativity Constraints

Constraints (2w) imply non-negativity restrictions on the number of individuals who are susceptible, being vaccinated, close contacts, successfully vaccinated, infected, total treated, recovered, funeral, buried, and treated, respectively, under scenario *ω*. Constraints (2x) represent the integer requirements on the number of type-*a* ETCs to be opened in region *r* at the end of period *j* under scenario *ω*. Additionally, if the number of infected individuals is less than 1 in a region *r*, the value of integer variable corresponding to opening an *a*-bed ETC is forced to be zero, and thus there will be no ETC opened in that region.

Constraint (2y) represents the binary variable 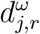, which is 1 when the number of infected individuals to be hospitalized is restricted by the number of available beds in the ETCs, and 0 when all infected individuals are hospitalized for treatment in the ETCs due to the available capacity.

## 5 Case Study and Results

### 5.1 Implementation Details

We apply our model 2a–2y to the case of the 2018-2020 EVD in the Democratic Republic of the Congo (DRC). The North Kivu and Ituri provinces of the DRC are affected by the EVD. We divided these two provinces into six different sub-regions: Upper (UNK), middle (MNK), and lower (LNK) North Kivu, and upper (UI), middle (MI), and lower (LI) Ituri. We describe the case study data used to formulate the model parameters, including population and migration data, resource cost data, and epidemiological data in Appendix B. The details of the mathematical formulation used to calculate the migration rates are presented in Appendix B.2.

We use a nested risk measure in multi-stage stochastic programming, known as the 𝔼-CVaR, in the stochastic model, as described in Section 3. We assume that the vaccines’ availability impacts the transmission rates in community contact, and thus uncertain transmission rates depend on the uncertain vaccine supply. Hence the high (low) availability of vaccines implies the low (high) realization of the uncertain transmission parameter. We present the value of the vaccine availability and uncertain transmission parameters under two realizations at each branch of the scenario tree, as shown in Table 10 in Appendix B.4. We solve the model for a 5-stage time period, where each stage corresponds to three weeks (from June 25, 2019, to October 8, 2019, in total 15 weeks) with high and low vaccine supply under different budget levels. Because each node of the scenario tree has two branches, each corresponding to a possible realization of the random parameters, we have 2^5^ = 32 scenarios for *T* = 5 stages.

The mathematical model is solved using CPLEX 12.7 on the desktop running with Intel *i*7 CPU and 64.0GB of memory. For each run, the time limitation is set at 72,000 CPU seconds. In the following subsections, we present results from solving the mean-risk multi-stage stochastic epidemics-vaccination-logistics model.

### 5.2 Resource Allocation under Different Budget Levels

We test the formulation for each time period under different budget levels: Very Tight ($40*M*), Tight ($70*M*), Medium ($100*M*), and Ample ($130*M*). The mean-risk trade-off coefficient *λ* is set to be 1, and the confidence level *α* is set to be 0.5.

Figure 2 presents the optimal allocation of resources (budget, capacity [number of beds], and vaccine) for each region under each budget level. According to the results, the regions with the highest initial infestation levels receive most of the budget, as in the case of Upper North Kivu and Middle North Kivu (Figure 2a). When the budget increases from very tight to ample, other regions will also get their share of the budget depending on their initial infection levels and disease-growth scenarios in the model.

**Figure 2:**
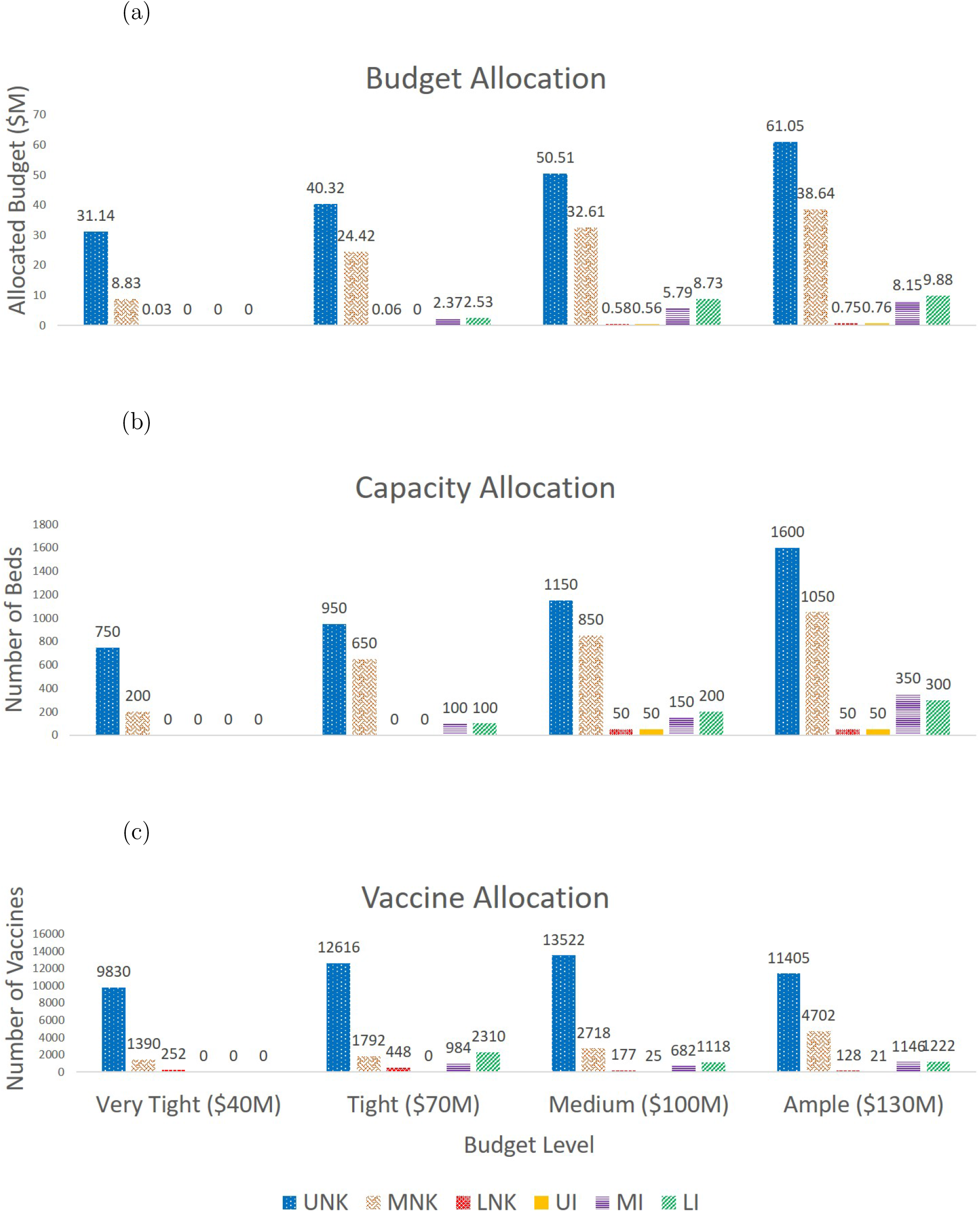
Budget, Capacity (Bed), and Vaccine Allocated to Each Region (*λ* = 1 and *α* = 0.5)

Similar to the total budget allocation, the total capacity is allocated based on the initial infection levels (Figure 2b). However, different from the total budget allocation, the majority of the ETCs are assigned in the first stage of the 5-stage planning horizon. For instance, under the $130*M* budget level, 1450 beds are allocated to Upper North Kivu in the first stage, while the total number of beds allocated to Upper North Kivu only increases by 150 by the end of stage five. Therefore, quick response in terms of allocating most of the treatment centers at the beginning of the outbreak will help to slow down the spread of the disease, while the majority of the budget should be allocated to treat infected people in ETCs and vaccine close contacts over multiple periods throughout the planning horizon.

When the budget level increases, the number of vaccines allocated to each region does not always increase (Figure 2c). For instance, under the $100*M* budget level, Lower North Kivu receives 177 vaccines, while this reduces to 128 vaccines under the $130*M* budget level. A possible reason for this is that vaccination can prevent more people from being infected, but it is not the most efficient way to reduce infections. Thus, the model gives priority to opening new ETCs and treating infected individuals and then uses the rest of the budget to allocate vaccines.

### 5.3 Analysis of the Risk Trade-Off

We perform an analysis of the risk parameters *λ* and *α* in terms of their impact on the objective function values and the resource allocation strategies. Specifically, under the $70*M* budget level, we compare four different problems with respect to their risk-averseness level, adjusting *λ* and *α* values accordingly–risk-neutral (*λ* = 0, *α* = 0), weak risk-aversion (*λ* = 1, *α* = 0.05), mild risk-aversion (*λ* = 10, *α* = 0.5) and strong risk-aversion (*λ* = 100, *α* = 0.95).

#### 5.3.1 Impact, Risk, and Cost under Risk-Neutral and Risk-Averse Policies

In this section, we provide insights on the effect of risk parameters *λ* and *α* on the expected impact and expected risk and how those values change as the decision maker shifts from being risk-neutral to risk-averse at varying risk levels. To analyze the results with the risk trade-off coefficient, we decompose the objective function into the *expected impact* [𝔼 (*f* (*x, ω*))] and the *expected risk* [*λ*CVaR_*α*_(*f* (*x, ω*))], as demonstrated in Equation (1). Specifically, the expected impact represents the expected value of the total number of infections, funerals, and close contacts of infected people, and the expected risk corresponds to the expected *CV aR*_*α*_ term in Eq. (2a). Table 1 presents results for the optimal objective function value for Eq. (2a), expected impact, expected risk, and the expected total cost over the four different risk-averseness levels described above. We also present values of the expected impact and expected risk for various combinations of *λ* = {0, 1, 10, 100} and *α* = {0.05, 0.5, 0.95} under the $70*M* budget level in Table 2. In both Tables 1 and 2, the “Expected Risk” under the risk-neutral model, where *λ* = 0, is computed as the expected total number of infections, funerals, and close contacts of infected people over all periods for the (1 − *α*) worst scenarios.

**Table 1:** Comparison of Objective Value, Expected Impact, Expected Risk, and Expected Cost under Various Risk-Averseness Levels

| | **Risk**<br>**Neutral**<br>($\lambda = 0, \alpha = 0.95$) | **Weak**<br>**Risk-aversion**<br>($\lambda = 1, \alpha = 0.05$) | **Mild**<br>**Risk-aversion**<br>($\lambda = 10, \alpha = 0.5$) | **Strong**<br>**Risk-aversion**<br>($\lambda = 100, \alpha = 0.95$) |
| --- | --- | --- | --- | --- |
| Objective Value <sup>a</sup> | 3,881,046 | 7,771,744 | 44,365,553 | 409,005,446 |
| Expected Impact | 3,881,046 | 3,881,501 | 3,882,344 | 3,884,815 |
| Expected Risk <sup>b</sup> | 5,628,125 | 3,890,243 | 4,048,321 | 4,051,206 |
| Expected Cost (\$M) | 69.70 | 69.76 | 69.84 | 69.85 |

<sup>a</sup> The calculation with units is represented as “Expected Impact” + $\lambda$ \* “Expected Risk”
<sup>b</sup> The Expected Risk is the $\text{CVaR}_\alpha$ and does not include preceding $\lambda$ coefficient

**Table 2:** Expected Impact and Expected Risk for Different Risk-averseness Levels

| $\lambda \backslash \alpha$ | 0.05 | | 0.5 | | 0.95 | |
| --- | --- | --- | --- | --- | --- | --- |
|  | Expected Impact | Expected Risk <sup>a</sup> | Expected Impact | Expected Risk <sup>a</sup> | Expected Impact | Expected Risk <sup>a</sup> |
| **0** | 3,881,046 | 3,957,443 | 3,881,046 | 4,512,163 | 3,881,046 | 5,628,125 |
| **1** | 3,881,501 | 3,890,243 | 3,881,323 | 4,047,577 | 3,887,508 | 4,054,319 |
| **10** | 3,881,053 | 3,889,802 | 3,882,344 | 4,048,321 | 3,883,418 | 4,050,082 |
| **100** | 3,883,462 | 3,892,219 | 3,881,704 | 4,047,882 | 3,884,815 | 4,051,206 |

<sup>a</sup> The Expected Risk is the $CVaR_\alpha$ and does not include preceding $\lambda$ coefficient

According to Table 1, the risk-neutral model has the smallest expected impact but the largest expected risk compared to all other risk-averseness levels. This indicates that the risk-neutral model provides the least expected number of infections, funerals, and close contacts over all scenarios, but once one of the worse-case scenarios happens, it gives a much larger number of infections, funerals, and close contacts compared to the risk-averse model. When both *λ* and *α* increase, the level of risk-averseness increases.Consequently, the optimal objective function value significantly increases due to the additional values of risk that are added to the objective formulation. The expected impact also increases, implying that there is a price for being risk-averse in terms of the increased number of infections, funerals, and close contacts under a more risk-averse policy. Moreover, the total cost of allocated capacity, treatment, and vaccination increases slightly when the risk-averseness level increases, implying that more budget is needed as the decision maker becomes more risk-averse.

Each row of Table 2 shows the change of the expected impact and expected risk when we fix the value of one of the risk parameters (*λ* = {1, 10, 100} and *α* = {0.05, 0.5, 0.95}) and change the other. All optimality gaps for the computational results in Table 2 are between 0.51% and 1.26%. According to the results, the largest expected risk occurs when *λ* = 0 for each *α* value. Similar to Table 1, in Table 2, the expected risk is the highest, and the expected impact is the lowest under the risk-neutral model. When we move from risk-neutral (*λ* = 0) to risk-averse (*λ* = {1, 10, 100}), the expected impact always increases. However, changing the *λ* parameter have a non-monotonous influence on the expected impact. When fixing the *λ* value and increasing the *α* value, the expected risk increases because we increase the confidence level for the impact of scenarios that is under the *V aR* value. When we increase *α* from 0.05 to 0.95, the risk-averseness level increases and the expected impact also shows an increasing trend for all risk-averse policies (*λ* ≥ 1).

#### 5.3.2 Resource Allocation under different Risk-Averse Policies

Table 3 presents the allocation of the budget over the five stages of the planning horizon. We observe that the majority of the budget is allocated under stages one, two, and four, while as the risk-averseness increases, the budget under stage one is slightly decreased and moved further into the future to be used later. The total budget used over all stages increases as the decision-maker becomes more risk-averse.

**Table 3:**
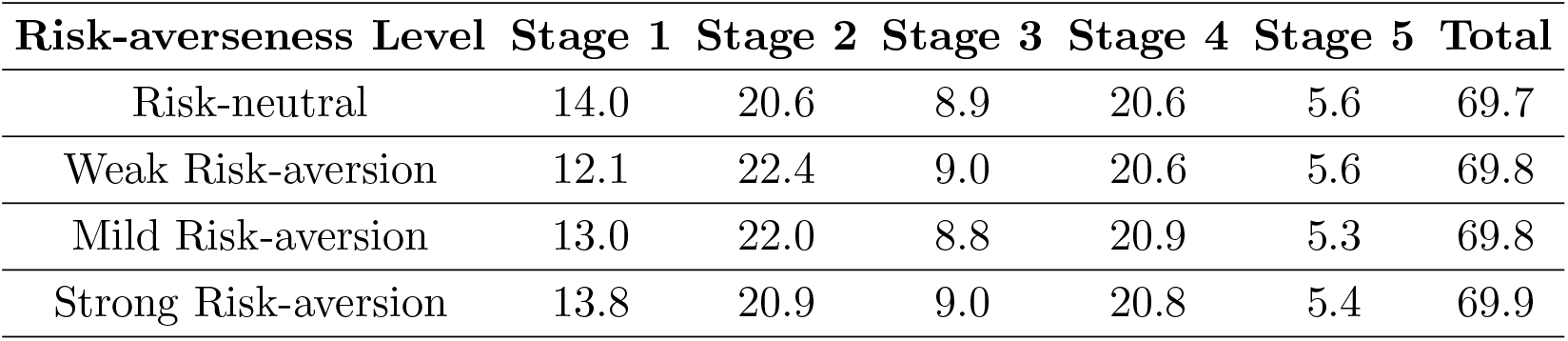
The Budget ($*M*) Allocation at Each Stage over Four Different Risk-Averseness Levels

Figure 3 illustrates the budget allocation of treatment, ETC capacity, and vaccination for different risk-averseness levels. As shown here, the capacity and treatment budget shows an increasing trend, and the budget allocated to vaccination shows a decreasing trend when the risk-averseness level increases. This may imply that as the decision-maker becomes more risk-averse, more investment is made on treatment rather than vaccination. It helps reduce variability in the set of scenarios that results in the highest number of infections and deaths.

**Figure 3:**
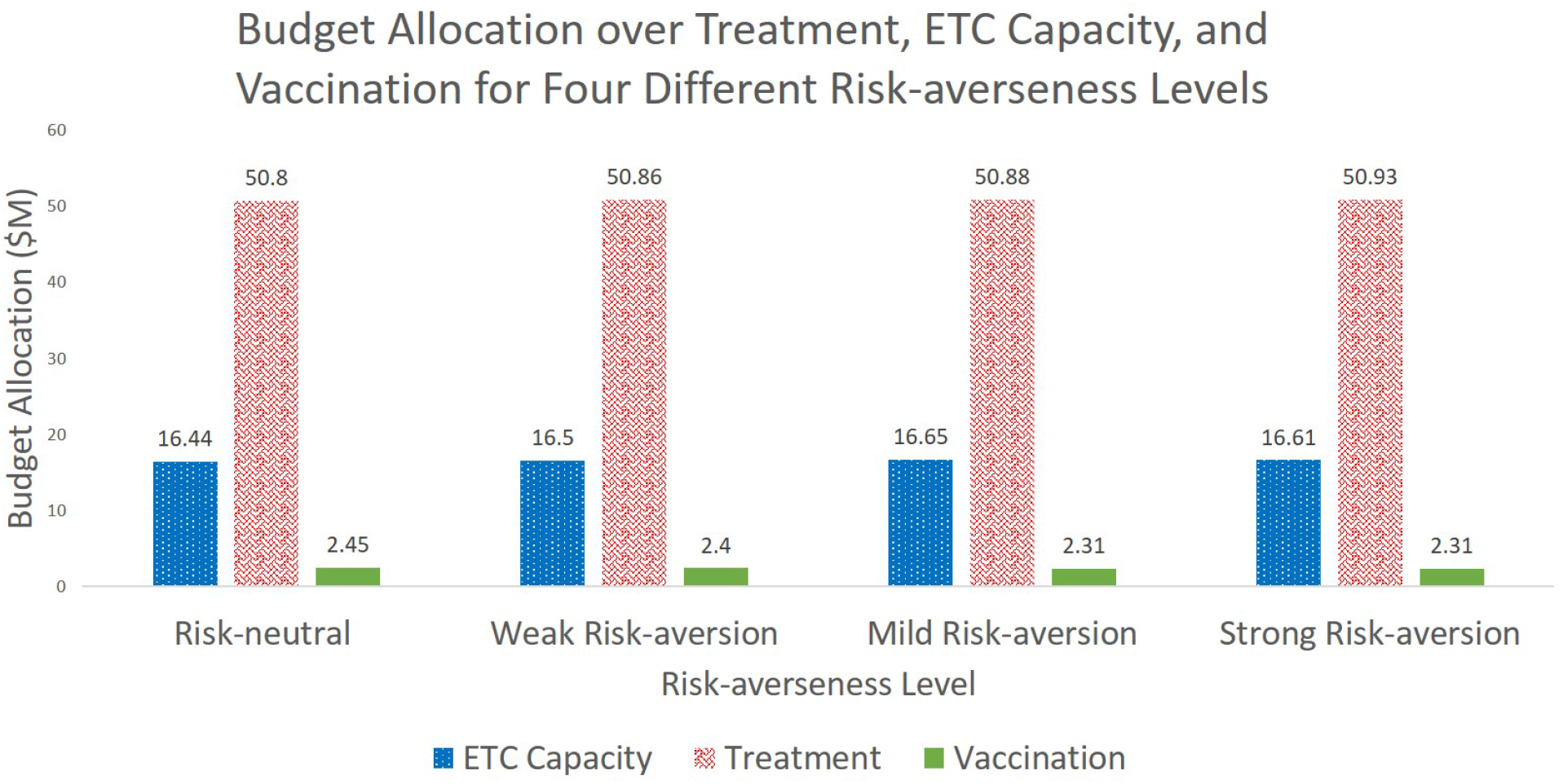
Budget Allocation over Treatment, ETC Capacity, and Vaccine for Four Different Risk-averseness

The optimal allocation of treatment budget, ETC capacity, and vaccine allocation under different risk-averseness levels is presented in Figure 4. When the risk-averseness level increases, the budget allocated to Upper North Kivu, which has the highest initial infection level, is decreased by moving the budget to neighboring locations with a potential risk of getting infections from Upper North Kivu (Figure 4a). For example, under the strong risk-averseness level, the budget allocated to Middle Ituri and Lower Ituri increases because they are geographically close to Upper North Kivu and thus are under the risk of getting infections through the large migrant population from Upper North Kivu.

**Figure 4:**
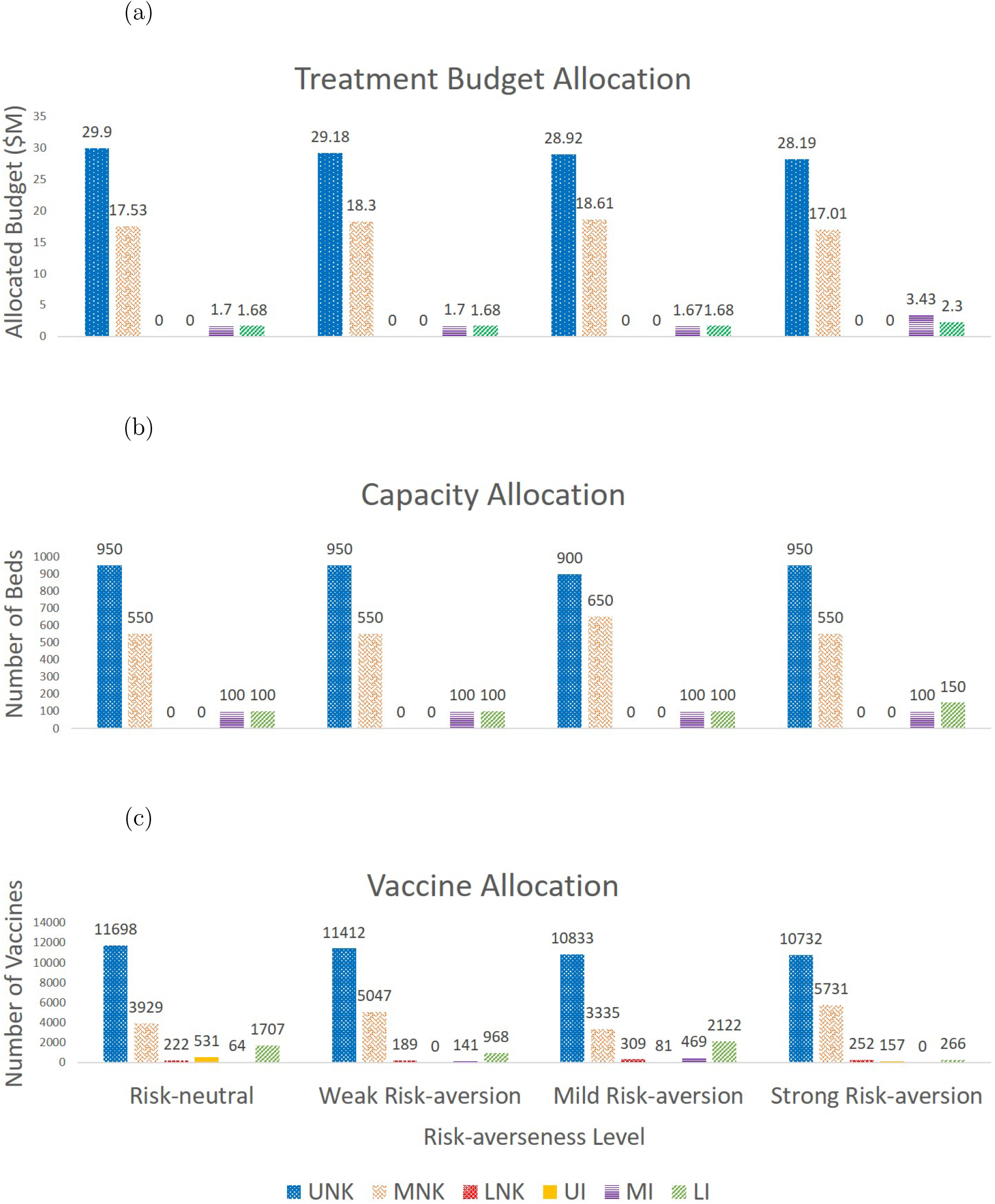
Treatment Budget, Capacity (Bed), and Vaccine Allocation under Different Risk-averseness Levels

The total capacity allocation of 1700 beds over all regions under the risk-neutral and weak risk-averseness cases is increased to 1750 beds under the mild and strong risk-averseness levels to alleviate adverse outcomes under the worst-case scenarios exceeding the VaR value (Figure 4b). As the risk-averseness level increases, the model allocates more beds to some regions that have low infection levels, considering their proximity to highly-infected locations and the risk of getting more infections from those locations. For example, Lower Ituri, right above the Upper North Kivu, receives an increased number of beds under the strong risk-averseness level.

The number of vaccines allocated to each region varies under different risk-averseness levels (Figure 4c). The total number of allocated vaccines decreases when the risk-averseness level increases. Regions that initially have zero infections get zero bed capacity. However, they still get vaccination due to the migration impact and the expectation of the disease spread under the risk-neutral and all risk-aversion cases. Thus, in locations where the disease has just started, the model uses the budget for vaccination to stop or slow the epidemic’s growth rather than building new ETCs.

### 5.4 Analyzing the Impact of Delay in Vaccination

In this section, we compare the number of infections and funerals for each stage for the 5-stage planning horizon with respect to the delay of the vaccine application in the first stage (Delay 1), first two stages (Delay 2), first three stages (Delay 3), first four stages (Delay 4) and five stages (Delay 5) under the $70*M* budget level. For example, in “Delay 3”, we fix the number of available vaccines in the first three stages to zero. In those first three stages, we assume a high transmission rate, which is equivalent to the low vaccine supply, and thus results presented here provide a lower bound on the number of infections and deaths. Here, we also consider a mild risk-averseness level by setting *λ* at 10 and *α* at 0.5.

The results of the cumulative number of infections and funerals in each type of delay in vaccines are shown in Figure 5, while Figure 6 demonstrates the non-cumulative infections and funerals for each delay option over no delay of vaccines for five stages. Both figures indicate that the number of infections and funerals increases exponentially over time for each delay type. The more the vaccination is delayed, the larger the infections and funerals are. Furthermore, the difference in the number of infections and funerals between “Delay 1” and “Delay 3” is larger than the difference between “Delay 3” and “Delay 5.” A possible reason is that the vaccination at the initial stages of the disease transmission would decrease the transmission rate faster and cause fewer infections and funerals than vaccination at later stages. These results indicate that the vaccines should be supplied as soon as possible once the epidemic breaks out, given the vaccines’ availability.

**Figure 5:**
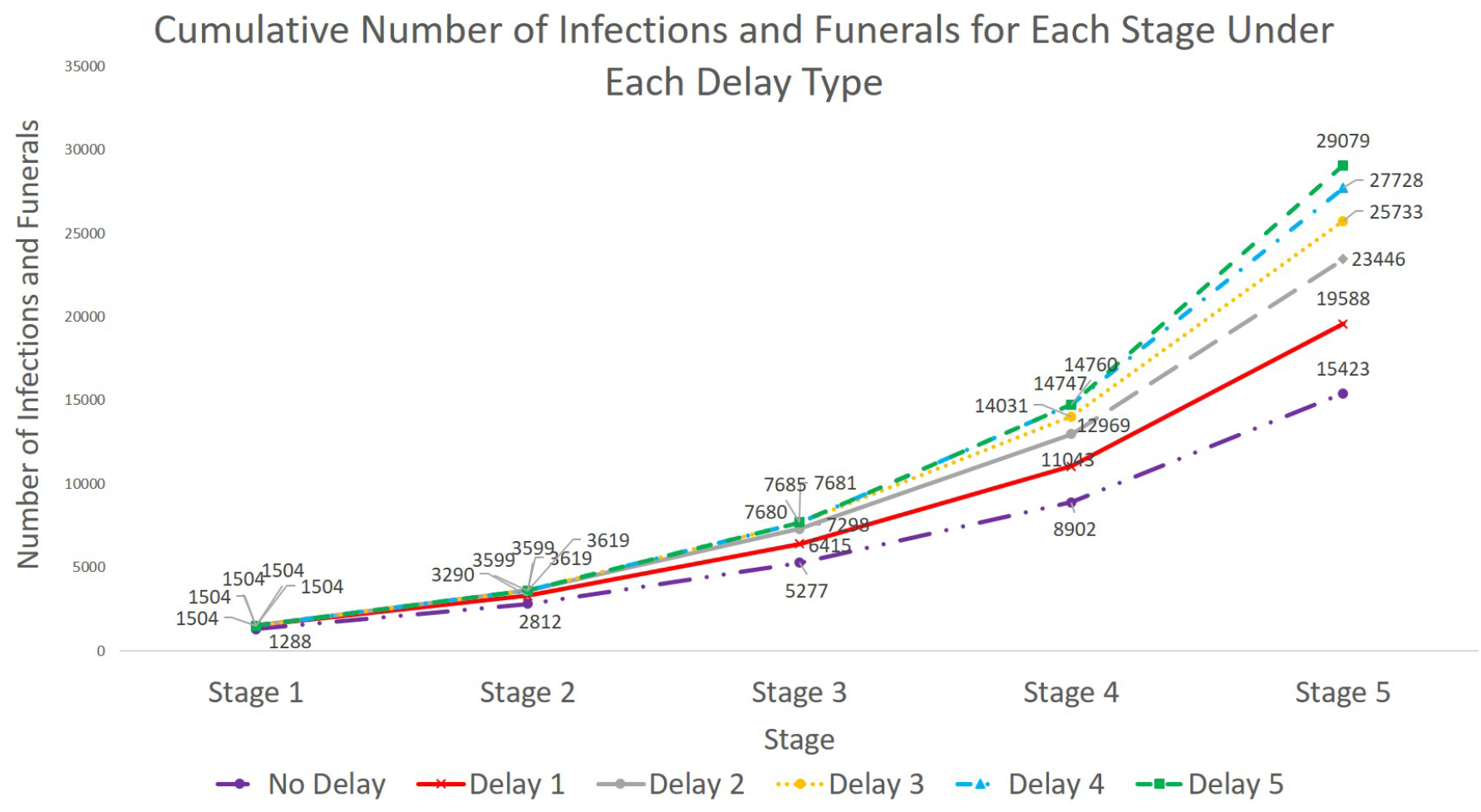
Cumulative Number of Infections and Funerals for Each Stage under Different Types of Delay (*λ* = 10 and *α* = 0.5)

**Figure 6:**
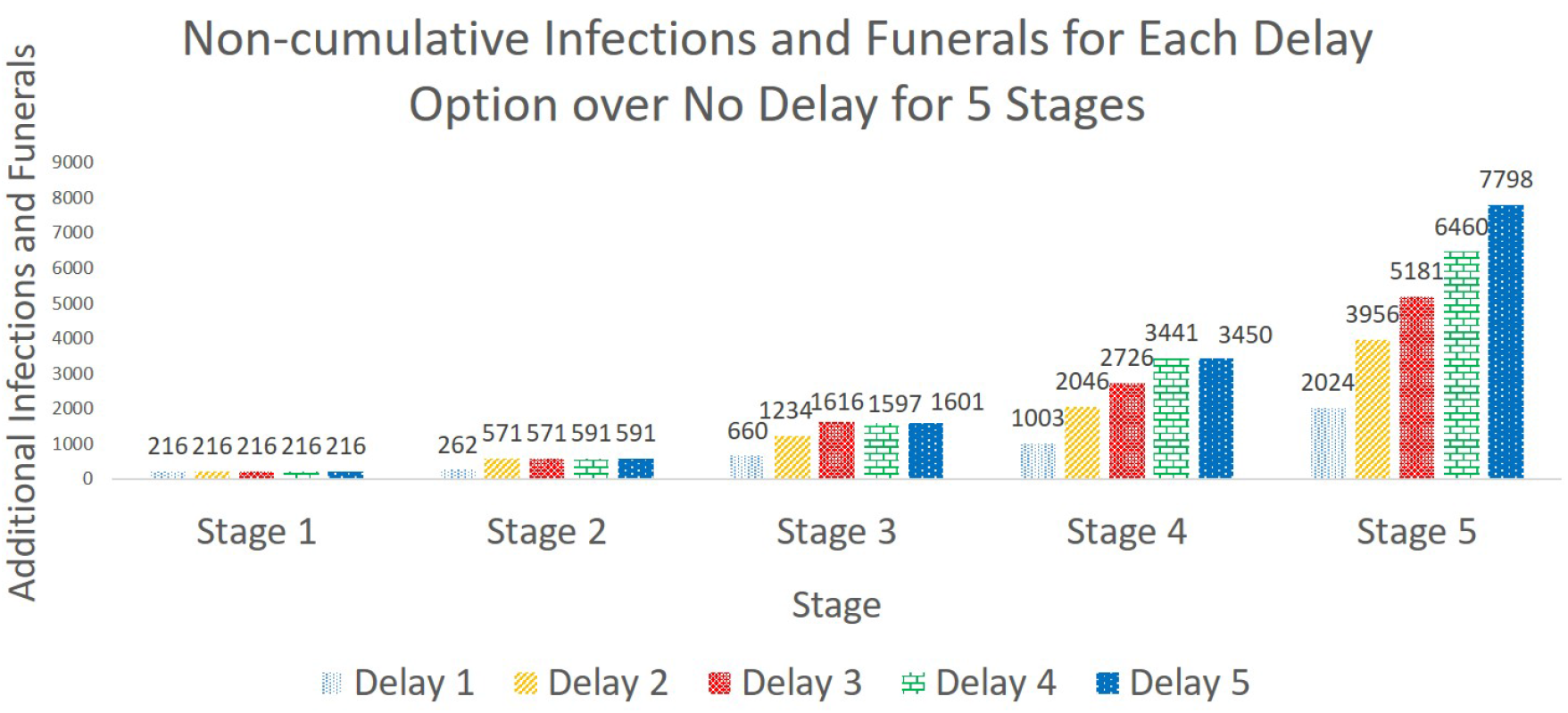
Non-cumulative Number of Infections and Funerals for Each Delay Option over No Delay (*λ* = 10 and *α* = 0.5)

## 6 Analyzing Vaccine Effectiveness and Acceptance Rates

In this section, we perform the sensitivity analysis on vaccination effectiveness and acceptance rates. Specifically, we adjust the vaccination effectiveness rate and the proportion of close contacts who are willing to get vaccinated. In WHO (2019a), the vaccination effectiveness rate is estimated as 0.975. In our model, we test vaccination effectiveness rates varying from 0.7 to 0.975 in increments of 0.05. Specifically, we change the value of the parameter *β*_*r*_ from low to high to simulate the impact of varying vaccination effectiveness rates on the number and location of vaccines allocated.

Several papers study the willingness of people to be vaccinated. For instance, Kpanake et al. (2018) find that 38% of people always choose to be immunized for the Ebola Virus Disease. Mudatsir et al. (2019) conclude that 74% of the people who participate in the interview express their acceptance of an Ebola vaccine. In addition, Ughasoro et al. (2015) suggest that 80% of the respondents accept being vaccinated with the Ebola vaccine. Therefore, we consider four different vaccine acceptance rates in the sensitivity analysis, which are 0.01, 0.4, 0.8, and 1. In reality, the vaccine acceptance rate may not be as low as 0.01. However, considering that the vaccine supply is quite limited, even under a vaccine acceptance rate of 0.4, the number of people who are willing to get vaccinated is still more than the upper bound on the vaccine supply. Thus, we use a vaccine acceptance rate of 0.01 to observe the changes in the vaccine allocation when the upper bound of vaccine supply is larger than the number of people who are willing to get vaccinated. In particular, we use a new parameter *f* to represent the number of close contacts who are willing to be vaccinated. Thus, Constraint (2r) is replaced by:

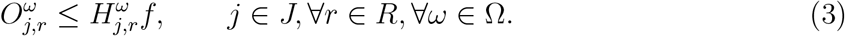

Constraint (3) implies that the number of successfully vaccinated individuals at stage *j* in region *r* under scenario *ω* should be less than or equal to the number of close contacts who are willing to be vaccinated.

In this section, we assume that the amount of available vaccine supply changes with respect to the time period. For example, in the initial stages of an epidemic or a vaccine discovery, it is less likely to make sufficient vaccine supply available to all demand locations. As time progresses, the vaccine supply upper bound will also increase. In this section’s analysis, the vaccine supply upper bound under each scenario is set to be 1000 at stage one, 2000 at stage two, 4000 at stage three, 5000 at stage four, and 6000 at stage five. The budget level for each test instance is set to be an ample level of $130*M*. Since the number of people who are willing to get vaccinated at the initial stages is less than the number of available vaccines, the remaining vaccines can be allocated in the following stages. Thus, in this analysis, Constraint (2s) is replaced by:

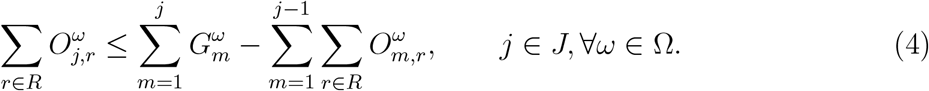

Constraint (4) ensures that the number of vaccines allocated to region *r* at stage *j* under scenario *ω* should be less than or equal to the total number of available vaccines up to stage *j* minus the total number of vaccines that are allocated to various regions from stage 1 until the end of stage *j* − 1.

Figure 7 shows the optimal number and location of vaccines allocated under different vaccination effectiveness rates when the vaccine acceptance rate is fixed at 0.8. For all the cases, the Upper North Kivu has most of the vaccines allocated, followed by Middle North Kivu. Also, the number of vaccines allocated to these two regions has a complementary relationship. When Upper North Kivu receives more (fewer) vaccines, Middle North Kivu will get fewer (more) vaccines allocated. This is because these two regions suffer from Ebola the most. The total number of infections and deaths does not fluctuate much if the number of vaccines allocated to these two regions is in a range of 11,000 to 15,000 for Upper North Kivu and 2,000 to 6,000 for Middle North Kivu. In addition, the total number of vaccines allocated under each case is close to the total supply upper bound of 18,000. Thus, we can conclude that, as long as the vaccine stays effective, no matter what vaccine effectiveness rate is, the total number of vaccines allocated will not significantly change under a limited supply of vaccines.

**Figure 7:**
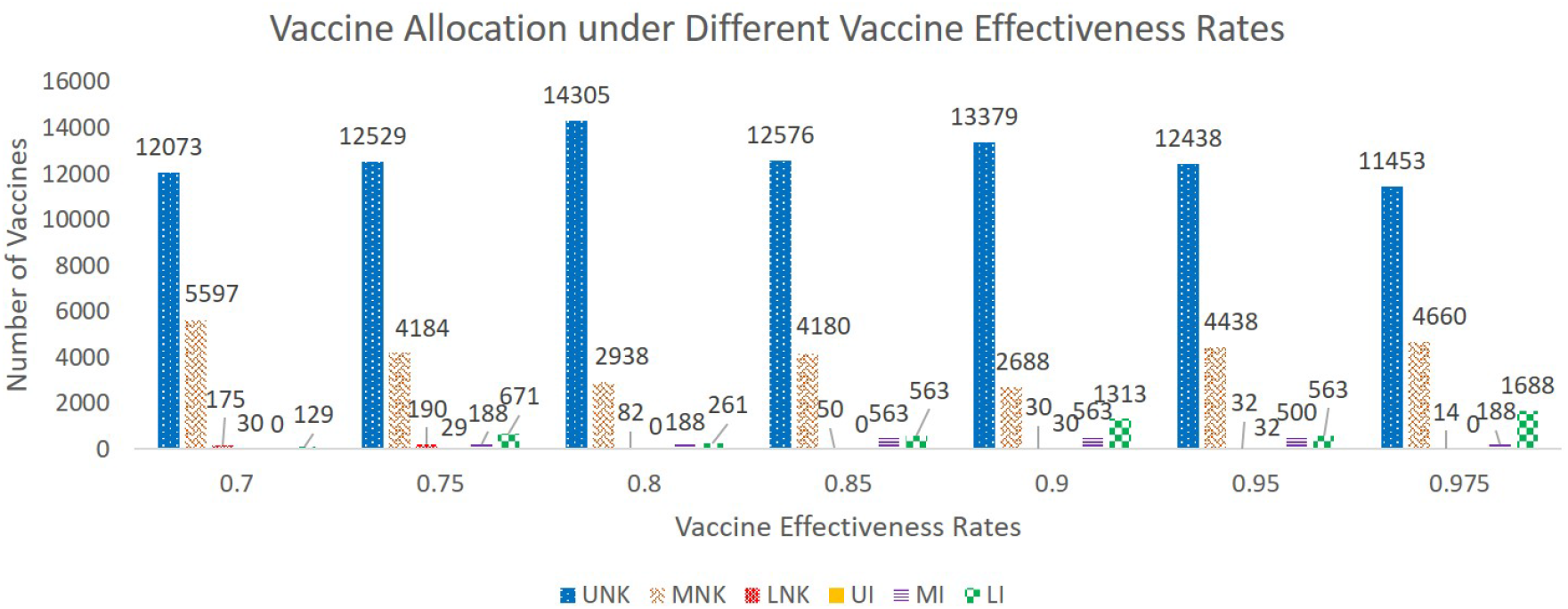
Vaccine Allocation under Different Vaccine Effectiveness Rates (*λ* = 10 and *α* = 0.95)

When the vaccination effectiveness rate increases, we also observe some of the vaccines that are allocated to regions with high levels of infections are moved to regions with fewer infections. Specifically, 17,670 vaccines allocated to Upper North Kivu and Middle North Kivu reduce to 16,113 vaccines when the vaccination effectiveness rate increases from 0.7 to 0.975. This result implies that the more effective the vaccine is, the fewer vaccines are needed in highly-impacted areas so that regions with lower infection could benefit from the remaining vaccines.

Figures 8a and 8b present the results for the first-stage and total vaccine allocation over all stages, respectively, under different vaccine acceptance rates when the vaccine effectiveness rate is fixed at 0.975. When the vaccine acceptance rate is 0.01, fewer vaccines are allocated in the first stage compared to the number of vaccines allocated under vaccine acceptance rates of 0.4, 0.8, and 1. Upper North Kivu is the only region that receives vaccines at the first stage when the vaccine acceptance rate equals 0.4, 0.8, and 1 because it has the highest number of initial infections among all regions. However, under the vaccine acceptance rate of 0.01, the number of close contacts willing to get vaccinated in Upper North Kivu is less than the supplied vaccines in the first stage. Thus, some vaccines in hand are allocated to other regions.

**Figure 8:**
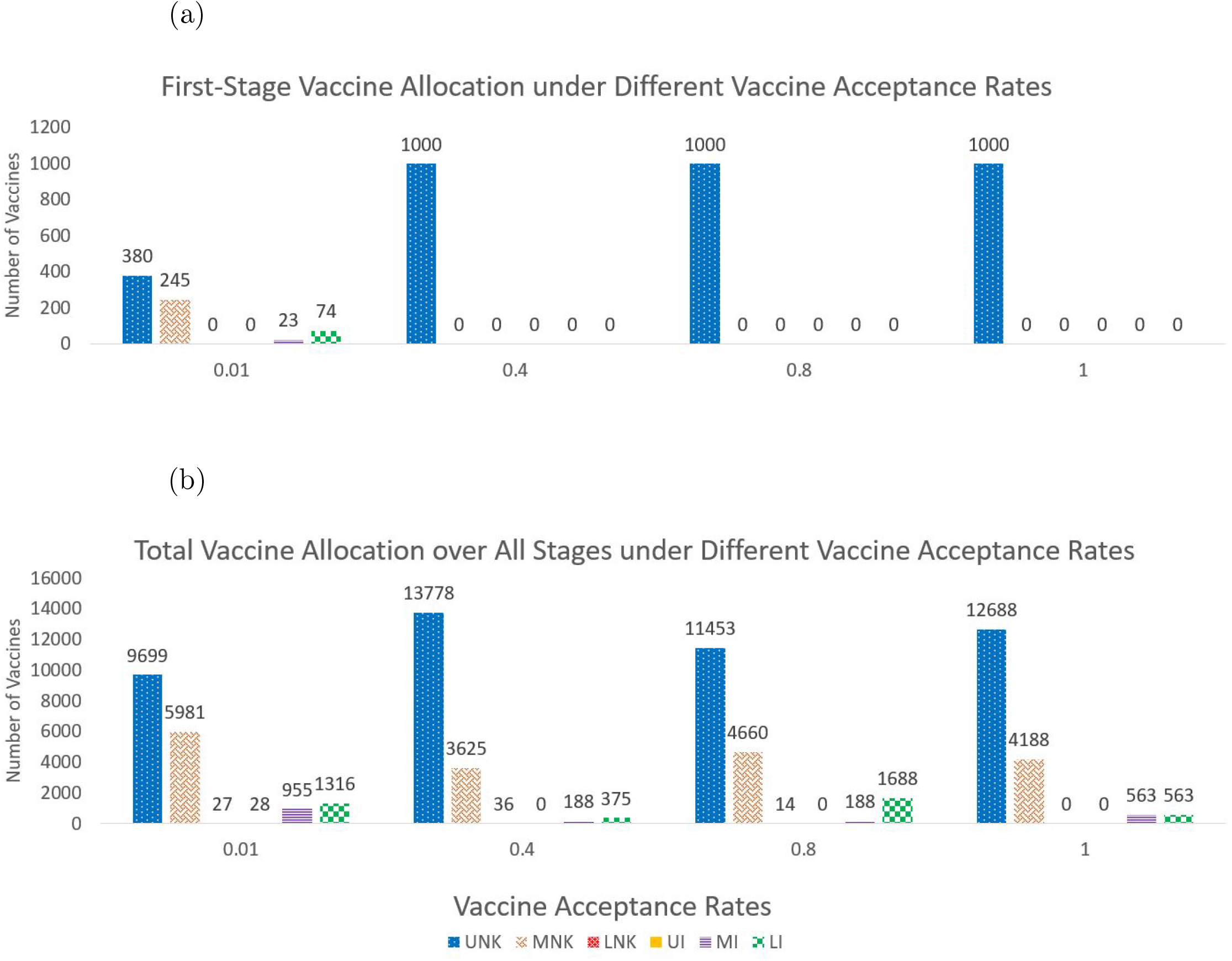
The First-Stage and Total Vaccine Allocation under Different Vaccine Acceptance Rates (*λ* = 10 and *α* = 0.95)

Compared to the first stage vaccine allocation, the total vaccine allocation over all stages shows a similar trend under different vaccine acceptance rates. This is because as the number of infections increases in the following stages, the number of close contacts also significantly increases. Therefore, even under the vaccine acceptance rate of 0.01, the number of close contacts who are willing to get vaccinated is more than the vaccine supply upper bound when considering the planning horizon of five stages. Furthermore, remaining vaccines from previous time stages will be used in the following stages. The total number of vaccines allocated to Upper North Kivu under the 0.01 vaccine acceptance rate is less than those in other cases. This is because more vaccines become available under a quite low vaccine acceptance rate, and those available vaccines are allocated to other regions with infections lower than that of Upper North Kivu.

## 7 Discussion and Conclusions

This paper presents a multi-stage mean-risk epidemics-vaccination-logistics model to address the optimal resource allocation challenges for epidemic control. We apply our model to the 2018-2020 EVD case in the Democratic Republic of the Congo (DRC). Because the information regarding the migration rates between regions in DRC is limited, we develop a method to estimate the transmission rate between each considered region. In our multi-stage stochastic programming model formulation, we use the CVaR in a nested form over multiple stages to minimize the total expected number of infections, funerals, and close contacts of infected people, and a weighted risk in a five-stage planning horizon, including 15 weeks.

The results regarding the analysis of risk trade-off show that there is a price for being prepared for the worst set of disease-growth scenarios. In other words, a risk-averse decision-maker should expect a possible increase in the number of infections and deaths while trying to mitigate disastrous outbreak scenarios. Thus, when the mean-risk trade-off coefficient increases, the confidence level in a risk-averse model will improve while the expected impact on infections and deaths worsens. Furthermore, the total cost of treatment and vaccination increases as the decision-maker becomes more risk-averse.

Allocating the resources fully based on the initial infection level will increase the risk of experiencing more infections and deaths in some disease scenarios. For example, the initial infection level of Lower Ituri is not high. Still, the model considers the possible adverse scenarios that may happen in Lower Ituri and allocates more resources to Lower Ituri under the case of strong risk-aversion. Thus, the potential risk associated with the disease growth in regions that have low initial infection levels but are in close proximity to hot spots of infection should also be considered when making risk-averse decisions on epidemics resource allocation.

The analysis of resource allocation under different budget levels indicates that the initial infection level is the key parameter that influences the budget and capacity allocation among each region. The regions with high initial infection levels get more resources, similar to the findings in Yin and Büyüktahtakın (2021). Different from the former literature, we study the budget allocation trade-off between treatment and vaccination while also accounting for the disease transmission dynamics. The results of vaccine allocation under different budget levels suggest that priority must be given to treat and isolate infected individuals in ETCs, while vaccination is supplementary to treatment. This has also been justified by the study of Kucharski et al. (2016), which states that ring vaccination might be insufficient to contain the outbreak if standard measures for controlling the transmission are not working, as in the EVD case in West Africa in early 2014. In another study, Kretzschmar et al. (2004) state that ring vaccination can contain the smallpox disease provided the intervention measures are very useful. Being risk-averse reinforces our findings on the resource-allocation priority of treatment over vaccination.

Our results also show that as the number of stages with no vaccines supplied increases, the number of infections, funerals, and close contacts exponentially increases even under other intervention measures, such as treatment and isolation. This result implies that vaccination is still quite effective when performed in addition to standard intervention measures, as shown in the studies of Kretzschmar et al. (2004), Ajelli et al. (2016), and Merler et al. (2016). Under the same length of delay, the delay of the vaccine at the early stages will cause more infections and deaths compared to the delay in the late stages. This proves that when the vaccines are limited but available, we should supply them as early as possible to minimize the number of infections at the beginning of an outbreak. Similarly, Wells et al. (2019) mention that even modest delays in initiating vaccination could noticeably degrade the impact of the epidemic control.

Interestingly, the model allocates vaccination to regions that get no treatment resources under a limited budget because it estimates a disease spread to these locations due to human mobility. Specifically, the model uses the budget for vaccination in regions where the disease has just started to curb the growth of the epidemic, while in regions with high initial infections, the model gives priority to build new ETCs and treat infected people over vaccination.

The sensitivity analysis on the vaccine effectiveness and acceptance rates indicates that the number of vaccines supplied to Upper North Kivu and Middle North Kivu has a complementary relationship. The number of vaccines provided to these two regions fluctuates under different vaccine effectiveness rates when the vaccine acceptance rate is fixed. This fluctuation does not influence the number of infections significantly.

When the vaccine effectiveness rate is fixed, vaccine acceptance rates affect vaccine allocation in the initial stages. Specifically, available vaccines are moved from highly impacted locations to less affected areas under a very low vaccine acceptance rate. In the early stages of an epidemic, there are fewer infections and close contacts, and the number of close contacts who are willing to get vaccinated is less than the number of vaccines supplied when the vaccine acceptance rate is very low. However, in the subsequent stages of an epidemic, the number of infections increases, thus significantly increasing close contacts. Therefore, even under a low vaccine acceptance rate, the number of close contacts who are willing to get vaccinated is much more than the available vaccine supply as the disease progresses over time. Consequently, those leftover and newly-supplied vaccines are allocated in the stages following the first stage to locations with both high and low infection levels. Thus, the total number of vaccines distributed over the whole planning horizon is similar under different vaccine acceptance rates.

This study leads to a number of important future directions. For example, the method we present for calculating the migration rate between each region could be further improved. Human behavior and social effects could be incorporated into the calculation of migration rates. For instance, Miami is not a metropolis compared to New York City. However, a substantial amount of people may travel to Miami for vacation. In our model, we only chose the metropolis of each region as the center of movement but did not consider other regions that may also have large short-term population migrations. This research could be extended by improving our estimation by considering more complicated environments to formulate the migration-estimation model.

In our epidemic-vaccination-logistics model, the term “logistics” represents the spatial and temporal allocation of ETCs, treatment budget used at ETCs as a function of *T* (treated) people, and the number of vaccines in region *r* at the end of period *j* under scenario *ω*. Here logistics do not involve details, such as distributing vaccines to certain locations within a region and its transportation specifics. A future extension of this work could investigate the transportation details of both vaccination and treatment resources, such as distributing those resources to specified locations within a region and its associated costs. Future work could also specify the transmission rate parameter, *θ*_1,*r*_, as a function of the vaccines allocated in region *r*, converting the model into a stochastic non-linear mixed-integer program. Both future directions would require the development of advanced solution algorithms, such as decomposition methods, global optimization, and cutting plane algorithms, to tackle the increased complexity of the mathematical model.

In this study, we present a general multi-stage mean-risk epidemics-vaccination-logistics model. This epidemic model could be adopted, for example, in the case of the COVID-19 pandemic. In such a model, the susceptible individuals can be divided into multiple sub-compartments based on their risk, demographics, and behaviors. For example, susceptible individuals can be divided into different sub-groups, such as people who wear masks and people under quarantine, with each group having a different infection rate, depending on the intervention measures applied. We can also analyze specific groups for vaccination, e.g., doctors, nurses, and volunteers to be injected. Increasing the number of compartments in a heterogenous-mixing epidemiological model will further complicate the epidemic-vaccination-logistics optimization model. Thus, the analysis of multiple specified groups can be studied in a more detailed agent-based simulation rather than a large-scale meta-population model as in our case.

For the pandemic control, we should also consider the international imported infections for the selected regions. Also, the travel patterns of people during the pandemic might influence the transmission of the disease. Therefore, our future research would present new mathematical models that will control a pandemic and help optimize resource allocation decisions.

## Data Availability

All data used is provided in the paper.

## Acknowledgments

We gratefully acknowledge the support of the National Science Foundation CAREER Award co-funded by the CBET/ENG Environmental Sustainability program and the Division of Mathematical Sciences in MPS/NSF under Grant No. CBET-1554018.

## Appendices

### A Notation

Sets and indices:

*J* : Set of time periods, 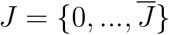.

*A* : Set of ETC types, *A* = {1, …, *Ā*}.

*R* : Set of regions, 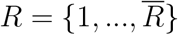.

*M*_*r*_ : Set of all surrounding regions of region *r*.

Ω : Set of scenarios, 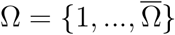.

*j* : Index for time period, where *j* ∈ *J*.

*r* : Index for region where *r* ∈ *R*.

*a* : Index defining type of ETC, where *a* ∈ *A*.

*ω* : Index for scenario, where *ω* ∈ Ω.

Transition parameters used to describe the rate of movement between disease compartments:

*χ*_1,*r*_ :Disease fatality rate without treatment in region *r*.

*χ*_2,*r*_ :Disease fatality rate while receiving treatment in region *r*.

*χ*_3,*r*_ :Disease survival rate without treatment in region *r*.

*χ*_4,*r*_ :Disease survival rate with treatment in region *r*.

*χ*_5,*r*_ :Safe burial rate of Ebola-related dead bodies in region *r*.

*σ*_*r*_ :Transmission rate per person in general community due to community interaction in region *r*.

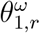:Transmission rate per person of close contacts due to interaction with infected individuals in region *r* under scenario *ω*.

*θ*_2,*r*_ :Transmission rate per person of close contacts during a traditional funeral ceremony in region *r*.

*f* :Vaccine acceptance rate.

*β*_*r*_ :Vaccine effectiveness rate.

*ε*_*r*_ :Transmission rate per person from successfully vaccinated V (immune) to general community S (not immune anymore) in region *r*.

Other parameters:

*b*_*j,r*_ :Unit cost of treatment for an infected individual in region *r* at the end of period *j*.

*g*_*aj,r*_ :Fixed cost of establishing type *a* ETC in region *r* at the end of period *j*.

*k*_*a*_ :Capacity (number of beds) of type *a* ETC.

*u*_*r*_ :The population in region *r*.

*e*_*j,r*_ :Unit cost per vaccine in region *r* at the end of period *j*.

Δ :Total available budget for treatment.

*π*_*r*_ :Initial number of susceptible individuals in general community in region *r*.

*φ*_*r*_ :Initial number of close contacts of infected people in region *r*.

*ϕ*_*r*_ :Initial number of vaccinated individuals in region *r*.

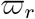:Initial number of infected individuals in region *r*.

*κ*_*r*_ :Initial number of treated individuals in region *r*.

*ϑ*_*r*_ :Initial number of recovered individuals in region *r*.

*υ*_*r*_ :Initial number of unburied dead bodies in region *r*.

*τ*_*r*_ :Initial number of buried dead bodies (funerals) in region *r*.

*ς*_*r*_ :Initial treatment capacity in terms of the number of ETC beds in region *r*.

*i*_*l*→*r*_ :Migration rate of infected individuals from surrounding regions *l* ∈ *M*_*r*_ to region *r*.

*h*_*l*→*r*_ :Migration rate of close contacts from surrounding regions *l* ∈ *M*_*r*_ to region *r*.

*i*_*r*→*l*_ :Migration rate of infected individuals from region *r* to surrounding regions *l* ∈ *M*_*r*_.

*h*_*r*→*l*_ :Migration rate of close contacts from region *r* to surrounding regions *l* ∈ *M*_*r*_.

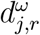:Binary variable for linearization in region *r* at the end of period *j* under scenario *ω*.

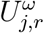:Auxiliary variable to be substituted with 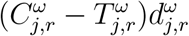 in region *r* at the end of period *j* under scenario *ω*.

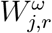:Auxiliary variable to be substituted with 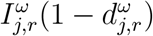 in region *r* at the end of period *j* under scenario *ω*.

*Q*_*LB*_ :Lower bound for 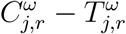.

*Q*_*UB*_ :Upper bound for 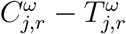.

*I*_*LB*_ :Lower bound for 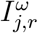.

*I*_*UB*_ :Upper bound for 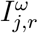.

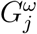:Number of total supplied vaccines at the end of period *j* under scenario *ω*.

*q* :Average number of close contacts per each infected individual.

State variables:

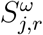:Number of susceptible individuals in general community in region *r* at the end of period *j* under scenario *ω*.

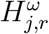:Number of close contacts of infected people in region *r* at the end of period *j* under scenario *ω*.

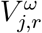:Number of successfully vaccinated individuals, who are fully immunized to disease, in region *r* at the end of period *j* under scenario *ω*.

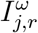:Number of infected individuals in region *r* at the end of period *j* under scenario *ω*.

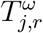:Number of individuals receiving treatment in region *r* at the end of period *j* under scenario *ω*.

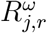:Number of recovered individuals in region *r* at the end of period j under scenario *ω*.

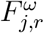:Number of deceased individuals due to the epidemic in region *r* at the end of period *j* under scenario *ω*.

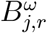:Number of buried individuals in region *r* at the end of period *j* under scenario *ω*.

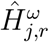:Number of close contacts of infected people migrating into region *r* at the end of period *j* under scenario *ω*.

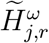:Number of close contacts of infected people emigrating from region *r* at the end of period *j* under scenario *ω*.

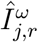:Number of infected individuals migrating into region *r* at the end of period *j* under scenario *ω*.

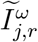:Number of infected individuals emigrating from region *r* at the end of period *j* under scenario *ω*.

Decision variables:

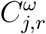:Total capacity (number of beds) in ETCs to be established in region *r* at the end of period *j* under scenario *ω*.

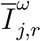:Number of infected individuals hospitalized (and quarantined) in region *r* at the end of period *j* under scenario *ω*.

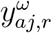:Number of type *a* ETCs established in region *r* at the end of period *j* under scenario *ω*.

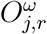:Number of vaccines allocated to region *r* at the end of period *j* under scenario *ω*.

Risk parameters:

*α* :Confidence level of value-at-risk, where *α* ∈ [0, 1).

*λ* :Non-negative risk preference parameter or mean-risk trade-off coefficient.

Risk variables:

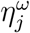:Value at risk at the confidence level *α* for each stage *j* under scenario *ω*.

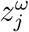:Value exceeding the value-at-risk at the confidence level 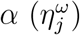 at stage *j* under scenario *ω*.

Sets and Parameters related to non-anticipativity:

*N* :Set of nodes in the scenario tree.

*n* :The serial number of nodes in the stochastic decision tree.

*β*(*n*) :Set of scenarios that pass through node *n* ∈ *N*.

*t*(*n*) :The corresponding stage that node *n* marked in the decision tree.

### B Ebola Case Study Data

This section presents the data used to formulate the model parameters, including population and migration data, resource cost data, and epidemiological data, and the mathematical formulation for estimating the migration rate.

#### B.1 Regional and Population Data

Based on the World Health Organization (WHO) Ebola situation report (WHO, 2020), we divided North Kivu and Ituri provinces of the Democratic Republic of Congo that are affected by the EVD into six different sub-regions: Upper (UNK), middle (MNK), lower (LNK) North Kivu and Upper (UI), middle (MI), lower (LI) Ituri. In the report of June 25th, 2019, there were almost no cases confirmed in Upper Ituri and Lower North Kivu; however, in the latest report of December 17th, 2019, some cases were confirmed in these regions. We consider the migration of people among multiple regions in our model and include these regions to analyze the influence of immigration on transmitting the disease.

Table 4 presents the population as well as the population ratio of each sub-region presented in Figure 9, while Table 5 shows the number of initial infections in each region on June 25, 2019.

**Table 4:** Regions, population size, and rate in West Africa

| North Kivu | Population<br>(millions) | Ratio | Ituri | Population<br>(millions) | Ratio |
| --- | --- | --- | --- | --- | --- |
| UNK | 1,9 | 0.19 | UI | 2,3 | 0.42 |
| MNK | 2,3 | 0.23 | MI | 2,8 | 0.51 |
| LNK | 5,7 | 0.58 | LI | 0,4 | 0.07 |
| **Total** | **9,9** | 1.00 |  | **5,5** | 1.00 |

**Figure 9:**
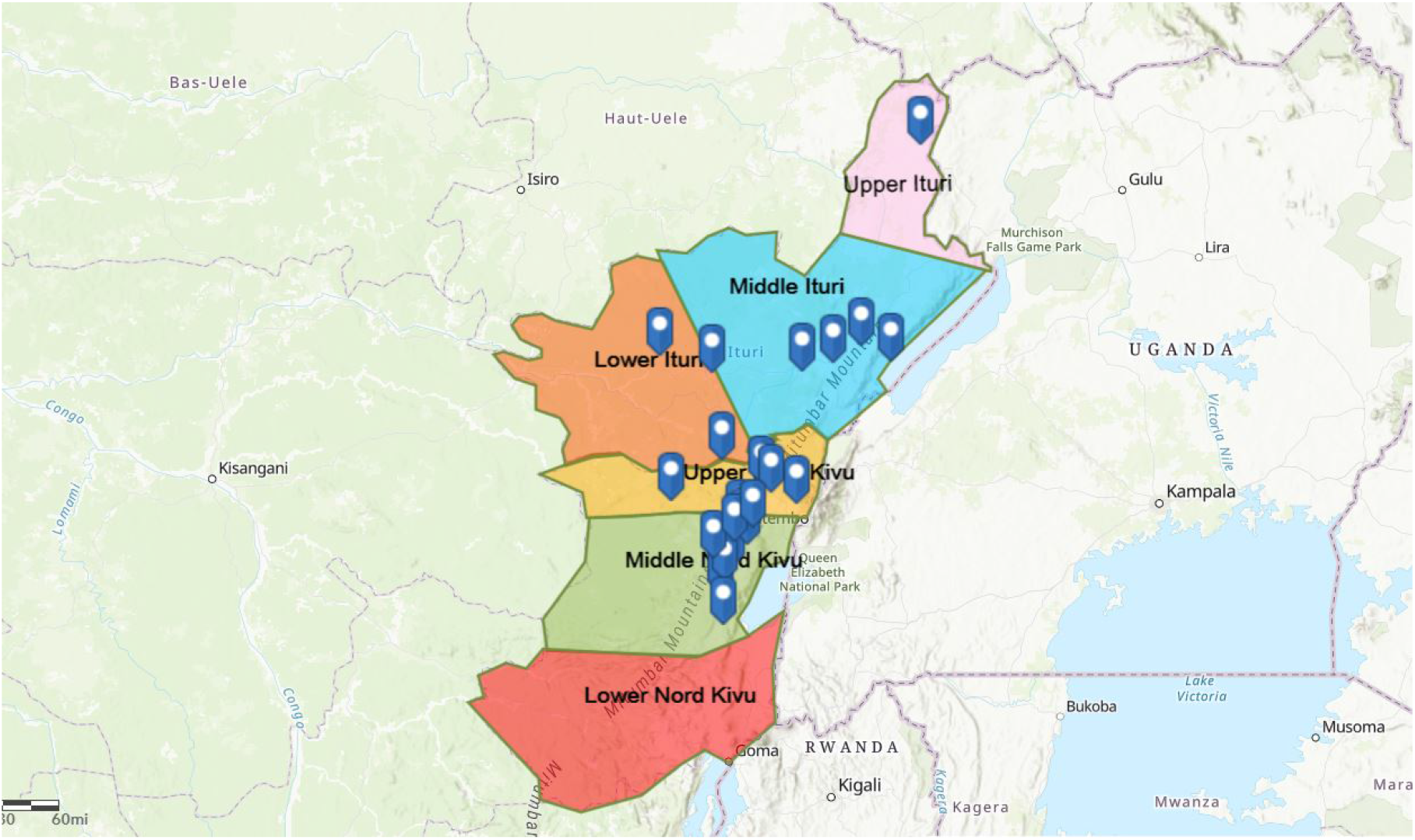
Region division in North Kivu and Ituri. The map is constructed using ArcGIS (2020).

**Table 5:** The number of infected people at the beginning of the planning horizon (June 25, 2019)

|  | Upper | Middle | Lower |
| --- | --- | --- | --- |
| North Kivu | 380 | 260 | 0 |
| Ituri | 0 | 23 | 74 |

#### B.2 Migration Estimation Model and Data for Migration Rates

Migration plays a crucial role in disease transmission between regions of a country and among multiple countries. Because the data regarding the migration of the population among each region of North Kivu and Ituri is not available, we have driven a new formula to estimate the rate of movement among multiple regions as described below. The logical sequence to derive the migration formulation is presented in Figure 10 and described below.

**Figure 10:**
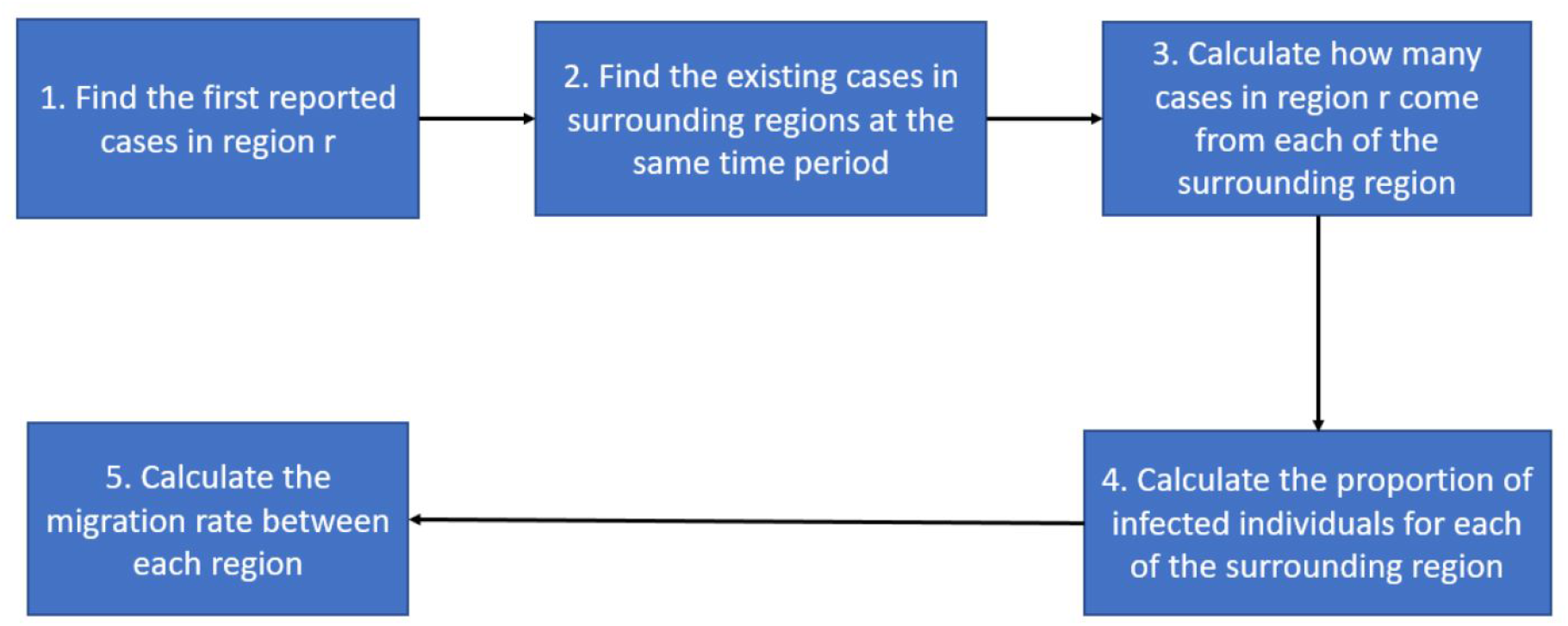
Steps of calculating the migration rate

If no border closure is imposed, some of the infected individuals from the regions where the disease has originated migrate to other regions during the incubation period of the virus. When disease symptoms begin to appear, they are discovered as the first cases in the new region. The Ebola virus average incubation period is around 2 to 21 days (WHO, 2019d). Thus, we estimate the migration rate using a cycle of three weeks, and the migration rate presented below is three-weekly. Let *r* ∈ *R* represents a newly infected region, and 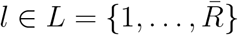 stands for the region where the disease has already existed, and 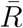 is the upper bound for the number of infected regions. Suppose we have a total of six regions as shown in Figure 9, *r* ∈ *R* = {*UNK, MNK, LNK, UI, MI, LI*}. For each region *r*, denote the number of infections detected in the first period (the first set of infected people) by *I*_*r*_.

##### 1. Find the First Reported Cases in Region *r*

We first check the WHO case situation report (WHO, 2020) and determine the first group of cases discovered in each defined region.

##### 2. Find Existing Cases in Surrounding Regions at the Same Time Period

We assume that the first group of infected individuals discovered in region *r* might have only come from the regions that already have infections at the same time period or before. Thus, using the WHO situation report (WHO, 2020), we choose a subset of regions *l* ∈ *L* surrounding region *r*, which have discovered infections before region *r*, as the possible emigration regions, where *I*_*l*_ ≠ 0. Specifically, since the average time from infected to either recovered and funeral, with or without treatment, is from one week to three weeks, we calculate the total confirmed cases in region *l* within the three-week interval of the time period where the first infection is seen in region *r, I*_*l*_.

##### 3. Calculate How Many Cases in Region *r* Might Have Come from Each of the Surrounding Region

In this step, we calculate the distance *D*_*rl*_ between each main region considered in Figure 9 using Google Maps (Table 6). We assume that the number of infected cases migrated from a possible emigration region *l* to region *r* is negatively correlated with *D*_*rl*_. Therefore, we compute the ratio of *D*_*rl*_ and the sum of the distances from region *r* to all possible emigration regions, and multiply it with the total migrated infected cases *I*_*r*_, to calculate the infected population in region *r* that resulted from people moving from region *l* to *r*, denoted by *Î*_*l*→*r*_, as given in below equation:

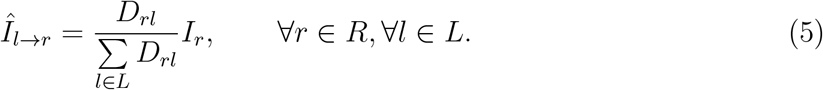

**Table 6:** Geographical distance between regions (KM)

| From \ To | UNK | MNK | LNK | UI | MI | LI |
| --- | --- | --- | --- | --- | --- | --- |
| UNK |  | 57.5 | 358 | 467 | 202 | 92.6 |
| MNK |  |  | 301 | 523 | 258 | 145 |
| MNK |  |  |  | 823 | 558 | 445 |
| UI |  |  |  |  | 267 | 483 |
| MI |  |  |  |  |  | 217 |
| LI |  |  |  |  |  |  |

##### 4. Calculate the Proportion of Infected Individuals for Each of the Surrounding Region

In this step, we calculate the infection ratio at region *l, i* = *I*_*l*_*/U*_*l*_, where *U*_*l*_ is the population of area *l*. We also assume that we should have the same ratio *i* for the people who immigrate from region *l* to region *r*. Then we have:

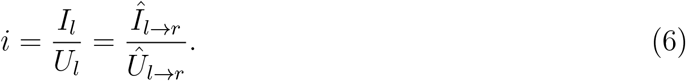

where *Î*_*l*→*r*_ represents the infected people who immigrate from region *l* to region *r*, as given in Equation (5), and *Û*_*l*→*r*_ stands for the migration population from *l* to *r*, which is the parameter that we want to estimate to calculate the migration rate.

##### 5. Calculate the Migration Rate Between Each Region

Substituting Equation (5) in Equation (6) and re-organizing it, the number of people that migrate from *l* to *r* can be calculated by the following equation:

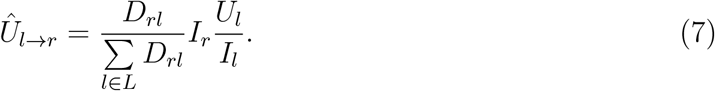

Since we formulate the model based on the short-term migration, we assume that those migration population is temporary (for business travel, cargo delivery, etc.) and migrated population will come back to the original region. Thus, the number of immigrated people from new infection regions *r* to the emigration regions *l* is estimated the same as the number of people moving from emigration regions *l* to the new infection regions *r*, i.e., *Û*_*l*→*r*_ = *Û*_*r*→*l*_.

Finally, the migration rate from region *l* to region *r* is calculated by dividing the immigration population from *l* to *r* with the total population of each region *l*, as given below:

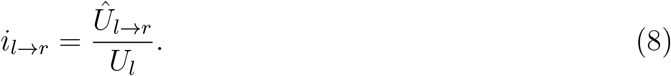

Because we have *Û*_*l*→*r*_ = *Û*_*r*→*l*_, similar to Equation (8), the migration rate from region *r* to region *l* is calculated by

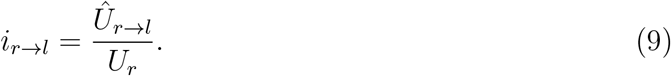

Additionally, suppose a considered region *b* ∈ *R* does not have any existed infections. In that case, we can estimate the migration rate based on the distance between region *l* and region *r*, for which the migration population from *l* to *r* is defined. Specifically, the migration between region *b* and region *r* can be estimated as:

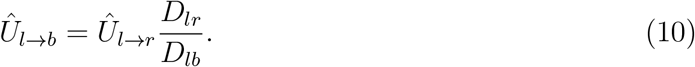

Then we can use Equations (8) and (9) to calculate the migration rate between region *b* and region *r*.

Here, we note that the estimation of the migration rate based on the distance between each region may not be accurate in some cases due to the popularity of a region and social effects. For instance, larger short-term migrations occur between cities rather than a city and a village because of the greater opportunities in urban locations and the corresponding social behavior. Therefore, to calculate the migration rates, the locations that have similar social environments should be selected from each region to minimize the impact of social effects on human mobility. In our case, each location selected from a region is a metropolitan of that region.

##### Example

As an example, consider the case of MNK. The WHO report (WHO, 2020) shows that MNK discovered the first three cases on September 11th, 2018, and at this time, only UNK and LI had infected cases. Thus, we assume that newly infected cases in MNK come from these two regions. The proportion of the distance from the main cities of UNK and LI to the main city in MNK is 57.5*/*145. So we assume that 71.6% (1.86) of the three cases in MNK come from UNK, and 29.4% (1.14) of them come from LI. The proportion of infections in UNK and LI is 0.00001 (21 over 1861730) and 0.00001 (2 over 148, 387). Using Equation (7), the migration population of UNK and LI to MNK is estimated as *Û*_*U NK*→*MNK*_ = 164, 896 and *Û*_*LI*→*M NK*_ = 84, 581. Then the migration rate from UNK to MNK is calculated as *i*_*UNK*→*MNK*_ = 0.0886, and the migration rate from LI to MNK is computed as *i*_*LI*→*MNK*_ = 0.5700. On the other hand, because we assume that *Û*_*M NK*→*UNK*_ = *Û*_*U NK*→*MNK*_ = 164, 896, the migration rate from MNK to UNK is calculated as *i*_*MNK*→*UNK*_ = 0.0685, and the migration rate from MNK to LI is given as *i*_*MNK*→*LI*_ = 0.0351.

The estimated migration rates between each region (Figure 9) are presented in Table 7.

**Table 7:** Migration rate between regions of North Kivu and Ituri.

| From \ To | UNK | MNK | LNK | UI | MI | LI |
| --- | --- | --- | --- | --- | --- | --- |
| UNK |  | 0.0886 | 0.0012 | 0.0016 | 0.0424 | 0.0345 |
| MNK | 0.0685 |  | 0.0059 | 0.0025 | 0.0851 | 0.0351 |
| LNK | 0.0081 | 0.0511 |  | 0.0935 | 0.0062 | 0.0023 |
| UI | 0.0138 | 0.0283 | 0.1200 |  | 0.0795 | 0.0088 |
| MI | 0.1254 | 0.0735 | 0.0050 | 0.0273 |  | 0.0167 |
| LI | 0.1456 | 0.5700 | 0.0044 | 0.0129 | 0.0711 |  |

#### B.3 Resource Allocation Cost Data

Table 8 gives the fixed cost of locating Ebola treatment centers (ETCs) and the variable or per-person cost of the Ebola treatment. The treatment cost includes the fixed cost for establishing each type of ETCs (either 50 or 100-bed ETC), isolation unit center, and laboratory diagnosis. Additionally, each facility has a variable running cost, mainly composed of treating infected people and contact tracing of the infected individuals. Safe burial cost is also included for safely burying infected dead bodies. Fixed costs are one-time; however, all other cost values given in Table 8 are presented for a three-week period. For example, an Ebola treatment center’s variable cost represents the cost of treating one infected individual over three weeks.

**Table 8:** Summary of fixed and variable treatment costs in 50 (100)-Bed ETC.

| Cost description | Fixed Cost | Variable cost* | Safe burial cost* |
| --- | --- | --- | --- |
| Ebola treatment center | \$386,000 (\$694,800) | \$13,215 | |
| Isolation unit center (IUC) | \$112,500 | \$1,699 | |
| Laboratory diagnosis | \$100,000 | \$810 | |
| Subnational technical services | | \$3,375 | |
| Contact tracing | | \$1,692 | |
| Safe burial | | | \$1,691 |
| **Total** | **\$598,500 (\$1,077,300)** | **\$20,791** | **\$1,691** |

\* Variable and safe burial costs are given for a period of three weeks.

**Table 9:** Transmission parameters and three-weekly rates for the Ebola outbreak.

| Parameter | Description | Data |  | Reference |
| --- | --- | --- | --- | --- |
|  |  | North Kivu | Ituri |  |
| $\chi_1$ | Rate of fatality without treatment | 0.527 | 0.372 | WHO (2019d), WHO (2014) |
| $\chi_2$ | Rate of fatality with treatment | 0.383 | 0.288 | WHO (2019d), WHO (2014) |
| $\chi_3$ | Rate of recovery without treatment | 0.695 | 0.725 | WHO (2019d), WHO (2014) |
| $\chi_4$ | Rate of recovery with treatment | 0.937 | 0.98 | WHO (2019d), WHO (2014) |
| $\chi_5$ | Safe burial rate | 2.22 | 2.13 | WHO (2019d), WHO (2014) |
| $\sigma_r$ | Transmission rate in the community | 0.648 | 0.54 | Camacho et al. (2014) |
| $\theta_{2,r}$ | Transmission rate at a traditional funeral | 2.04 | 2.19 | Camacho et al. (2014) |
| $\beta_r$ | Vaccination effectiveness rate | 0.975 | 0.975 | WHO (2019a) |
| $\varepsilon_r$ | Probability of transition from vaccinated to susceptibles | 0.06 | 0.06 | WHO (2018) |

**Table 10:**
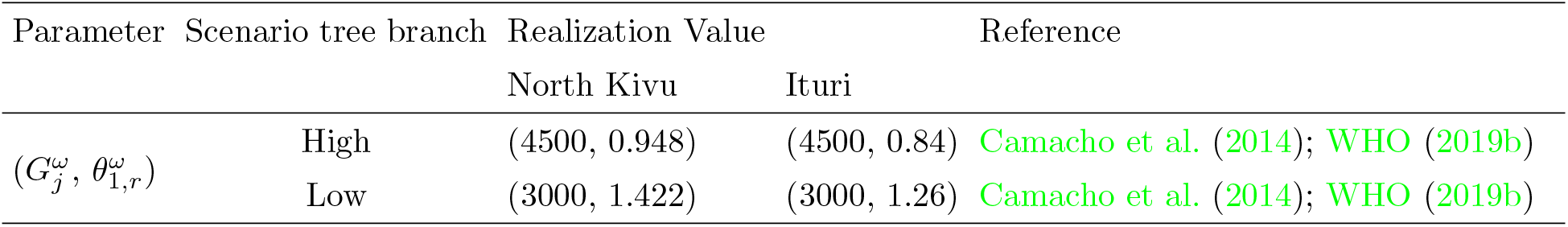
Three-weekly values for vaccine supply upper-bound 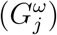 and the uncertain transmission rate 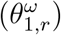 for the Ebola outbreak. High (low) realization for 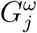 implies low (high) realization for 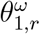.

#### B.4 Epidemiological Data

In this section, we present the values of the parameters that describe the disease transmission among compartments of the EVD that were described in Section 4.1 in North Kivu and Ituri of DRC.

